# T cell and antibody functional correlates of severe COVID-19

**DOI:** 10.1101/2020.11.25.20235150

**Authors:** Krystle K.Q. Yu, Stephanie Fischinger, Malisa T. Smith, Caroline Atyeo, Deniz Cizmeci, Caitlin R. Wolf, Erik D. Layton, Jennifer K. Logue, Melissa S. Aguilar, Kiel Shuey, Carolin Loos, Jingyou Yu, Nicholas Franko, Robert Y. Choi, Anna Wald, Dan H. Barouch, David M. Koelle, Douglas Lauffenburger, Helen Y. Chu, Galit Alter, Chetan Seshadri

**Affiliations:** Department of Medicine, University of Washington School of Medicine, Seattle, WA, USA; Ragon Institute of MGH, MIT and Harvard, Boston, MA, USA; PhD program in Immunology and Virology, University of Duisburg-Essen, Essen, Germany; PhD program in Virology, Division of Medical Sciences, Harvard University, Boston, MA, USA; Department of Biological Engineering, Massachusetts Institute of Technology, Cambridge, MA, USA; Center for Virology and Vaccine Research, Beth Israel Deaconess Medical Center, Harvard Medical School, Boston, MA 02215, USA; Providence Medical Group, Everett, WA, USA; Department of Epidemiology, University of Washington School of Public Health, Seattle, WA, USA; Department of Laboratory Medicine and Pathology, University of Washington School of Medicine, Seattle, WA, USA; Vaccine and Infectious Diseases Division, Fred Hutchinson Cancer Research Center, Seattle, WA, USA; Department of Global Health, University of Washington, Seattle, WA, USA; Benaroya Research Institute, Seattle, WA, USA

## Abstract

Comorbid medical illnesses, such as obesity and diabetes, are associated with more severe COVID-19, hospitalization, and death. However, the role of the immune system in mediating these clinical outcomes has not been determined. We used multi-parameter flow cytometry and systems serology to comprehensively profile the functions of T cells and antibodies targeting spike, nucleocapsid, and envelope proteins in a convalescent cohort of COVID-19 subjects who were either hospitalized (n=20) or not hospitalized (n=40). To avoid confounding, subjects were matched by age, sex, ethnicity, and date of symptom onset. Surprisingly, we found that the magnitude and functional breadth of virus-specific CD4 T cell and antibody responses were consistently higher among hospitalized subjects, particularly those with medical comorbidities. However, an integrated analysis identified more coordination between polyfunctional CD4 T-cells and antibodies targeting the S1 domain of spike among subjects that were not hospitalized. These data reveal a functionally diverse and coordinated response between T cells and antibodies targeting SARS-CoV-2 which is reduced in the presence of comorbid illnesses that are known risk factors for severe COVID-19. Our data suggest that isolated measurements of the magnitudes of spike-specific immune responses are likely insufficient to anticipate vaccine efficacy in high-risk populations.

## INTRODUCTION

Severe Acute Respiratory Syndrome Coronavirus 2 (SARS-CoV-2) causes Coronavirus Disease 2019 (COVID-19), which is responsible for over one million deaths since its discovery in early 2020 (1, 2). The clinical course of COVID-19 is variable and ranges from asymptomatic or mild disease to acute respiratory distress syndrome (ARDS) and death(3). Epidemiologic studies have revealed several factors, such as advanced age, male sex, and non-white ethnicity (4–6), that are associated with adverse clinical outcomes, including hospitalization. The presence of medical comorbidities, such as obesity, diabetes, and heart disease are also associated with more severe disease (7–9). Viral load at diagnosis is an independent predictor of mortality, and duration of viral shedding was longer among hospitalized patients who died (10, 11).

Several studies have identified lymphopenia and an increase in pro-inflammatory cytokines associated with hospitalization for COVID-19 (12–15). Neutralizing antibody titers have also been associated with increased disease severity (16–18). The role of the adaptive immune system in promoting immune pathology is further supported by autopsy studies, which have revealed the presence of infiltrating B and T lymphocytes in the heart and lungs of patients who died (19, 20). These data suggest that immune-mediated damage, as well as direct viral cytopathic effects, may be responsible for poor clinical outcomes after SARS-CoV-2 infection.

Detailed studies using flow and mass cytometry as well as single cell RNA sequencing have revealed perturbations in several sub-populations of T cells and B cells among patients with severe COVID-19 (21–25). However, T cells and antibodies execute a range of functions only after encountering their cognate antigens, so further details regarding their role in the pathogenesis of COVID-19 has required looking beyond bulk populations of lymphocytes. Several studies have investigated whether cross-reactive T cell and humoral responses are present in unexposed blood donors (26–29). However, these studies have not comprehensively examined the functions of antigen-specific T cells and have not been designed to robustly examine associations with clinical risk factors and outcomes. A major limitation has been confounding due to demographic factors, such as age and sex, as well as the date of symptom onset, all of which can influence associations with immune status independent of COVID-19 (30, 31). For example, some studies reported differences between acutely ill patients and healthy controls and recovered donors, but the healthy controls were significantly younger, and recovered donors had blood drawn much later in their illness course (21, 32).

In this study, we sought to overcome these limitations in study design and to more comprehensively examine the functional profiles of antigen-specific immune responses and their association with risk factors and clinical outcomes after COVID-19. We leveraged a large cohort of convalescent donors, including individuals recruited as candidate donors for convalescent plasma donation (33) in Seattle, WA, where SARS-CoV-2 community transmission was first described in the United States (34). We selected study participants that were either hospitalized (n=20) or not hospitalized (n=40) after matching for age, sex, ethnicity, and date of symptom onset. Archived serum was used to compare neutralizing antibody titers as well as immunoglobulin (Ig) levels, Fc receptor (FcR) binding, and Fc effector functions targeting full spike (S), S1, S2, receptor binding domain (RBD), and nucleocapsid (N) proteins. Archived peripheral blood mononuclear cells (PBMC) were used to compare frequencies and phenotypes of conventional αβ T cells as well as donor-unrestricted T cells (DURTs)(35). Finally, we compared the functional profiles of antigen-specific T cells targeting S1, S2, N, and envelope (E) proteins using intracellular cytokine staining (ICS). In nearly all the parameters tested, we consistently observed both higher magnitudes and increased functional breadth among hospitalized subjects, particularly those with medical comorbidities. However, T cell and antibody responses showed less correlation among hospitalized subjects. Our balanced analysis reveals a qualitative shift in the adaptive immune response to SARS-CoV-2, which may be directly related to the presence of comorbid illnesses that are known risk factors for severe disease.

## RESULTS

### Cellular and humoral dynamics in a matched cohort of convalescent COVID-19 subjects

We utilized a cohort of convalescent COVID-19 subjects stratified by hospitalization status and matched for confounders most relevant for immune profiling studies, namely age, sex, and ethnicity (Table 1). We further matched for the interval between the self-reported date of symptom onset and specimen collection, as this could also influence kinetics of SARS-CoV-2 specific immune responses(36). This resulted in a final set of COVID-19 subjects who were either hospitalized (n=20) or not hospitalized (n=40) and from whom plasma and peripheral blood mononuclear cells were collected within a median of ∼50 days post symptom onset (Table 1). Quantitative viral load information was available from 16 subjects and varied over a wide range (Supplementary Table 1). Consistent with prior reports, comorbid diseases were more frequently observed among hospitalized subjects (p=0.001, Fisher’s exact test) (7–9).

**Table 1.** Summary of demographics of the SARS-CoV-2 Convalescent Cohort. Study participants included COVID-19 subjects who were either hospitalized (n=20) or not hospitalized (n=40). The two groups were matched for age, sex, ethnicity, and date of symptom onset. Comorbid illnesses are indicated and were either self-reported or else abstracted from the patient’s electronic medical record. These include, but are not limited to, those presented in the table.

|  | **Non-hospitalized**<br>**(n = 40)** | **Hospitalized**<br>**(n = 20)** |
| --- | --- | --- |
| **Age (median, range), years** | 56.5 (23.0-79.0) | 59.0 (28.0-74.0) |
| **Sex (n, %)** |  |  |
| Female | 20 (50.0) | 8 (40.0) |
| **Race/ethnicity (n, %)** |  |  |
| White, non-Hispanic/Latino | 34 (85.0) | 13 (65.0) |
| African American, non-Hispanic/Latino | 0 (0.0) | 0 (0.0) |
| Other, non-Hispanic/Latino | 5 (12.5) | 6 (30.0) |
| Hispanic/Latino | 1 (2.5) | 1 (5.0) |
| **Insurance Status (n, %)** |  |  |
| Public (or none/self-pay) | 6 (15.0) | 4 (20.0) |
| Private or both | 33 (82.5) | 8 (40.0) |
| Unknown | 1 (2.5) | 8 (40.0) |
| **Days Since Symptom Onset (median, range)** | 49.5 (26.0-74.0) | 54.5 (32.0-71.0) |
| **Comorbidities (n, %)** |  |  |
| Diabetes | 2 (5.0) | 5 (25.0) |
| Heart Disease | 0 (0.0) | 2 (10.0) |
| Hypertension | 6 (15.0) | 4 (20.0) |
| Other | 1 (2.5) | 6 (30.0) |

We used multi-parameter flow cytometry and system serology to comprehensively study the functional profiles of T cells and antibodies targeting SARS-CoV-2 spike, nucleocapsid, and envelope proteins (Figure 1A). We also examined the neutralization activity of patient sera and noted consistent titers up to 74 days in this cross-sectional analysis. However, neutralization titers were not associated with hospitalization status (Figure 1B). This result suggested that other humoral or T cell functional profiles may be associated with clinical outcomes in COVID-19 subjects. We examined the magnitude of Ig subclasses targeting the full spike protein (S), the S1, S2 or receptor binding domain (RBD) of spike, and nucleocapsid (N), which were broadly stable in both groups of subjects over time (Supplementary Figure 1). IgG1, IgG2, IgG4, and IgA titers against full spike, S1, S2, and RBD were significantly higher among hospitalized subjects (Figure 1C and Supplementary Figure 2). Moreover, all Ig subclasses except IgG4 targeting nucleocapsid were also significantly higher among hospitalized subjects, and we have previously demonstrated that anti-nucleocapsid antibodies are a marker of disease severity (Figure 1C)(37). These results show that antibody subclass titers rather than neutralization may be associated with clinical outcomes after COVID-19.

**Figure 1.**
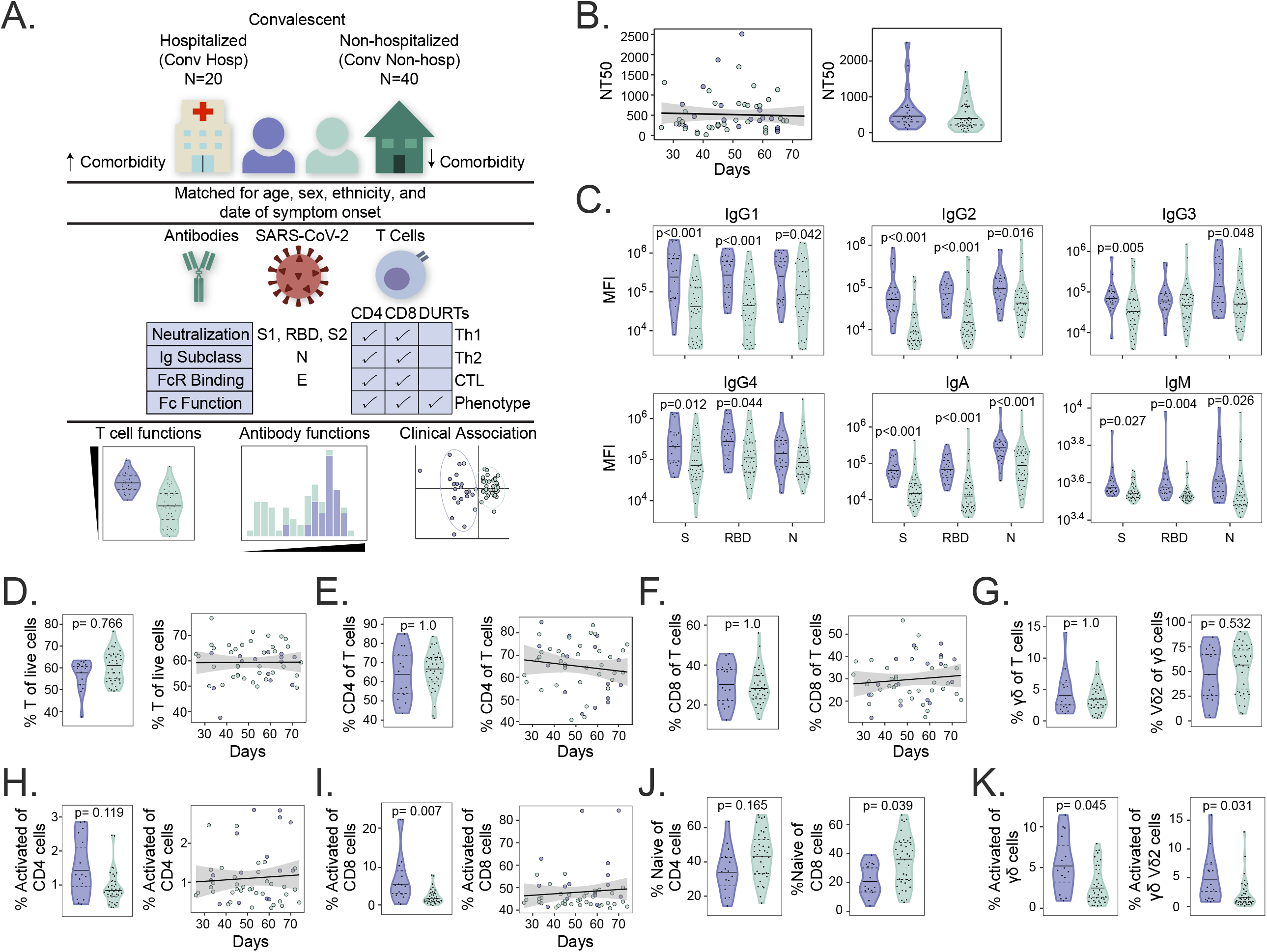
Cellular and humoral dynamics in a matched cohort of convalescent COVID-19 subjects. (A) Study schema. Archived peripheral blood mononuclear cells (PBMC) and plasma from COVID-19 study subjects that were previously hospitalized (purple, n=20) or non-hospitalized (green, n=40) were selected based on matching for age, sex, ethnicity, and date of symptom onset. Samples were comprehensively profiled for SARS-CoV-2 specific T cell and antibody phenotypes and functions. Data were analyzed to identify differences between the groups and to build a classifier. DURTs = Donor-unrestricted T cells (B) Antibody neutralization titers were compared between hospitalized and non-hospitalized subjects (left) and graphed according to days since symptom onset (right). NT50 denotes the concentration of serum required to achieve 50% of the maximum neutralization in the assay. (C) Comparison of antibody subclass and isotype levels against spike (S), receptor binding domain (RBD), and nucleocapsid (N) antigens between groups. (D) Flow cytometric analysis comparing the percent of total CD3+ T cells between groups and graphed according to days since symptom onset. Among CD3+ T cells, the percent of (E) CD4+ T cells and (F) CD8+ T cells was compared between groups and graphed according to days since symptom onset. (G) The frequency of γδ T cells are as a percent of total CD3+ T cells, and Vδ2 T cell frequencies as a percent of γδ T cells are compared between groups. The frequency of activated (H) CD4+ and (I) CD8+ T cells defined by co-expression of HLA-DR and CD38 are compared between groups and graphed according to date of symptom onset. (J) The percentage of naive CD4+ and CD8+ T cells as defined by co-expression of CD45RA and CCR7 is compared between groups. (K) The frequencies of HLA-DR+CD38+ γδ and Vδ2 T cells are compared between groups. NT50, Ig titers, and T cell frequencies were compared between groups using Mann-Whitney U tests, followed by correction for multiple hypothesis testing using the Bonferroni method. Median, 25th, and 75th quartiles are indicated for violin plots. Black lines on scatter plots represent the best fit linear regression line, and grey-shaded areas represent the 95% confidence interval of the predicted mean. If not shown, p-values for Mann-Whitney tests and regressions were not significantly different.

**Figure 2.**
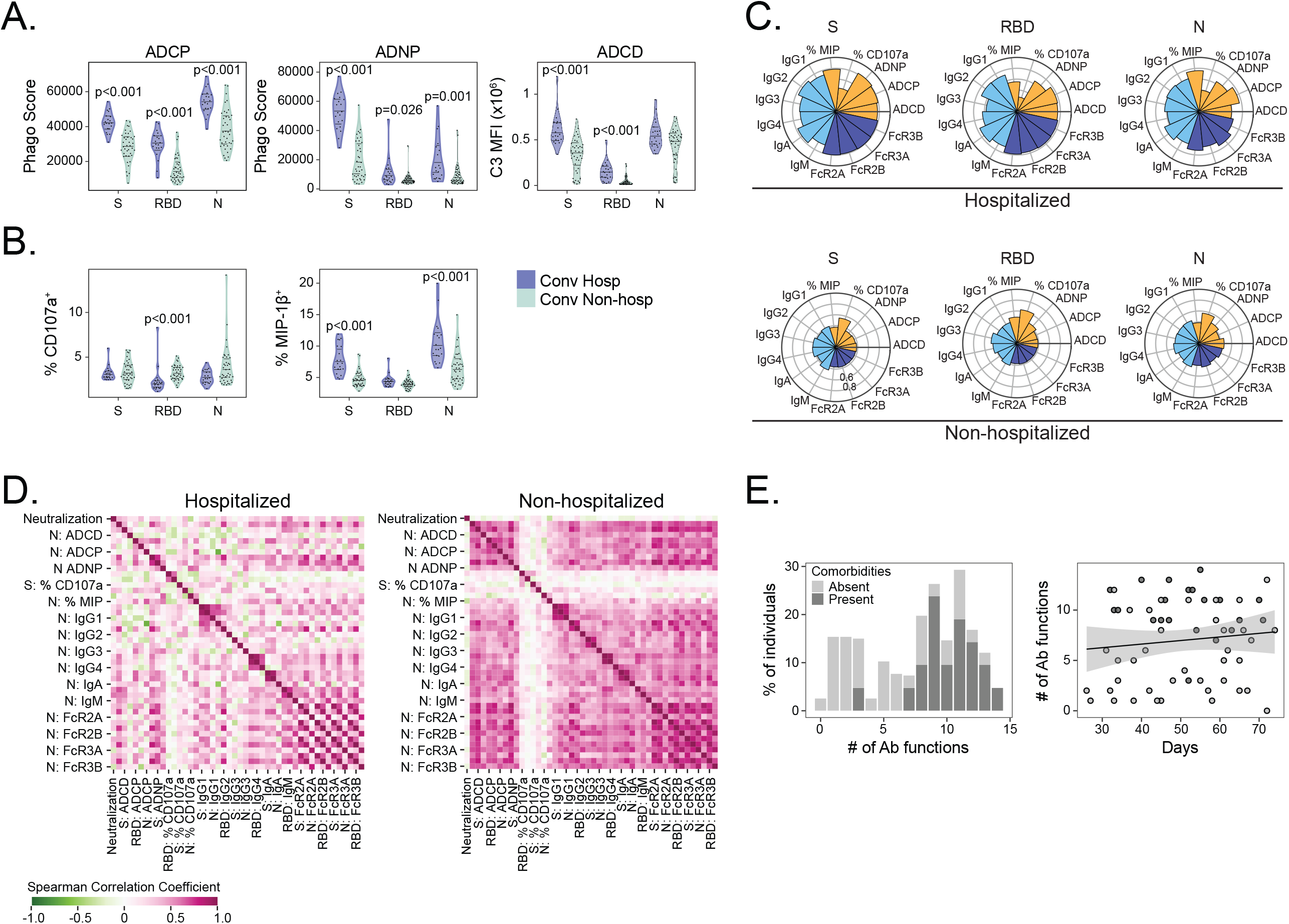
Antibody functional profiles are associated with hospitalization after COVID-19. SARS-CoV2-2 specific antibody phenotypes and functional profiles were compared between hospitalized (purple, n=20) and non-hospitalized (green, n=40) COVID-19 study subjects. (A) Antibody dependent cellular phagocytosis (ADCP), antibody dependent neutrophil phagocytosis (ADNP), antibody dependent complement deposition (ADNP) and (B) NK cell activation as measured by MIP-1β secretion or CD107a expression against spike (S), receptor binding domain (RBD), and nucleocapsid (N) was quantified and compared between groups. (C) Nightingale rose graphs show the distribution around the mean profiles of antibody features for S, RBD, and N among hospitalized and non-hospitalized subjects. Each flower petal represents a SARS-CoV-2 specific antibody measurement. The size of the petal depicts the percentile above/below the mean across both groups. The colors indicate type of feature: antibody function (orange), titer (light blue) and Fc-receptor binding (dark blue). (D) The correlation matrix shows the Spearman correlation coefficient for antibody features separately in subjects with and without comorbidities. Pink indicates a positive correlation, whereas green indicates a negative correlation. (E) Polyfunctional antibody profiles were compared between subjects with and without comorbidities. To determine polyfunctionality, an individual’s response was noted to be functional if it was above the median response for the cohort. Per person, the number of positive functions were summed, resulting in a polyfunctionality score per individual. Polyfunctional scores are displayed as percent positivity of the whole cohort. Antibody phenotypes and effector functions excluding neutralization were compared across cohorts using Mann-Whitney U tests followed by correction for multiple hypothesis testing using the Bonferroni method. Median, 25th, and 75th quartiles are indicated for violin plots. The black line on the scatter plot represents the best fit linear regression line, and the grey-shaded area represents the 95% confidence interval of the predicted mean. If not shown, p-values for Mann-Whitney tests and regression were not significantly different.

### Antibody functional profiles are associated with hospitalization after COVID-19

To follow up these differences in Ig subclass, we examined several Fc-binding specificities and Fc-dependent effector functions. Fc-receptors (FcRs) specificities FcR2A, FcR2B, FcR3A, and FcR3B binding full spike, S1, S2, RBD, and N were significantly higher among hospitalized subjects (Supplementary Figure 2). Antibody-dependent cellular phagocytosis (ADCP), antibody-dependent neutrophil phagocytosis (ADNP), and antibody-dependent complement deposition (ADCP) against full spike, RBD, and N was significantly increased among hospitalized subjects (Figure 2A). Notably, while MIP-1β secretion by natural killer (NK) cells was increased among hospitalized subjects, NK cell degranulation measured by CD107a expression was elevated among non-hospitalized subjects (Figure 2B). To obtain a qualitative summary of the differences in antigen-specific humoral responses between groups, we visualized Ig subclass, Fc-binding specificity, and Fc-effector functions targeting S, RBD and N using nightingale rose graphs (Figure 2C). The results show consistently higher levels of measured analytes among hospitalized subjects, with the exception of CD107a expression on NK cells. We next examined the correlation of antibody profiles independently in hospitalized and non-hospitalized subjects. The correlation with neutralization titers in both groups was low, supporting our analysis of non-redundant aspects of the SARS-CoV-2 specific antibody response. Relative to non-hospitalized subjects, hospitalized subjects demonstrated lower correlation among antibody titers, Fc-specificities, and Fc-effector functions (Figure 2D). This difference was robust to sub-sampling in order to account for the unequal sample sizes in each group (Supplementary Figure 7). Finally, we calculated a polyfunctionality score for each individual for S, RBD and N over the six antibody functionality readouts against three SARS-CoV-2 antigens. Subjects with comorbidities were able to activate a robust polyfunctional antibody response against S, RBD, and N in comparison to subjects without comorbidities (Figure 2E). Taken together, these results reveal qualitative and quantitative increases in several aspects of the SARS-CoV-2 specific antibody response among hospitalized subjects with comorbidities, many of which are likely the result of differences in innate immune system activation and T cell help.

### Activated CD8 and γδ T cells are associated with hospitalization after COVID-19

To investigate the role of T cells, we used multi-parameter flow cytometry to quantify the frequencies and phenotypes of conventional and donor-unrestricted T cell populations, such as invariant NK T (iNKT) cells, mucosal-associated invariant T (MAIT) cells, and γδ T cells(35). In our matched cross-sectional analysis, we noted that the frequency of CD3+, CD4+, and CD8+ T cells did not vary significantly over time since symptom onset or between hospitalized and non-hospitalized subjects (Figure 1D, 1E, 1F and Supplementary Figure 3). We also found no difference in the frequency of γδ T cells, invariant NKT cells, or mucosal associated invariant T cells as well as B cells, monocytes, or NK cells (Figure 1G and Supplementary Figure 4). However, the frequency of activated CD8+ T cells was significantly higher among hospitalized subjects, which is consistent with prior reports (Figure 1I) (21, 38, 39). The frequency of naive CD8+ T cells was also lower among hospitalized subjects, suggesting differentiation to an effector phenotype after SARS-CoV-2 infection (Figure 1J). Among total γδ T cells, the frequency of activated γδ T cells was higher among hospitalized subjects independent of expression of the Vδ2 gene segment (Figure 1K). The frequency of activated CD4, CD8, and γδ T cells was broadly steady over time since symptom onset, which is in contrast to some reports (Figure 1H, 1I, and Supplementary Figure 4C)(38). These data confirm and extend published studies by revealing the durability of differences in activated CD8 and γδ T cell but not CD4 T cell populations in a matched cross-sectional analysis stratified by hospitalization status.

**Figure 3.**
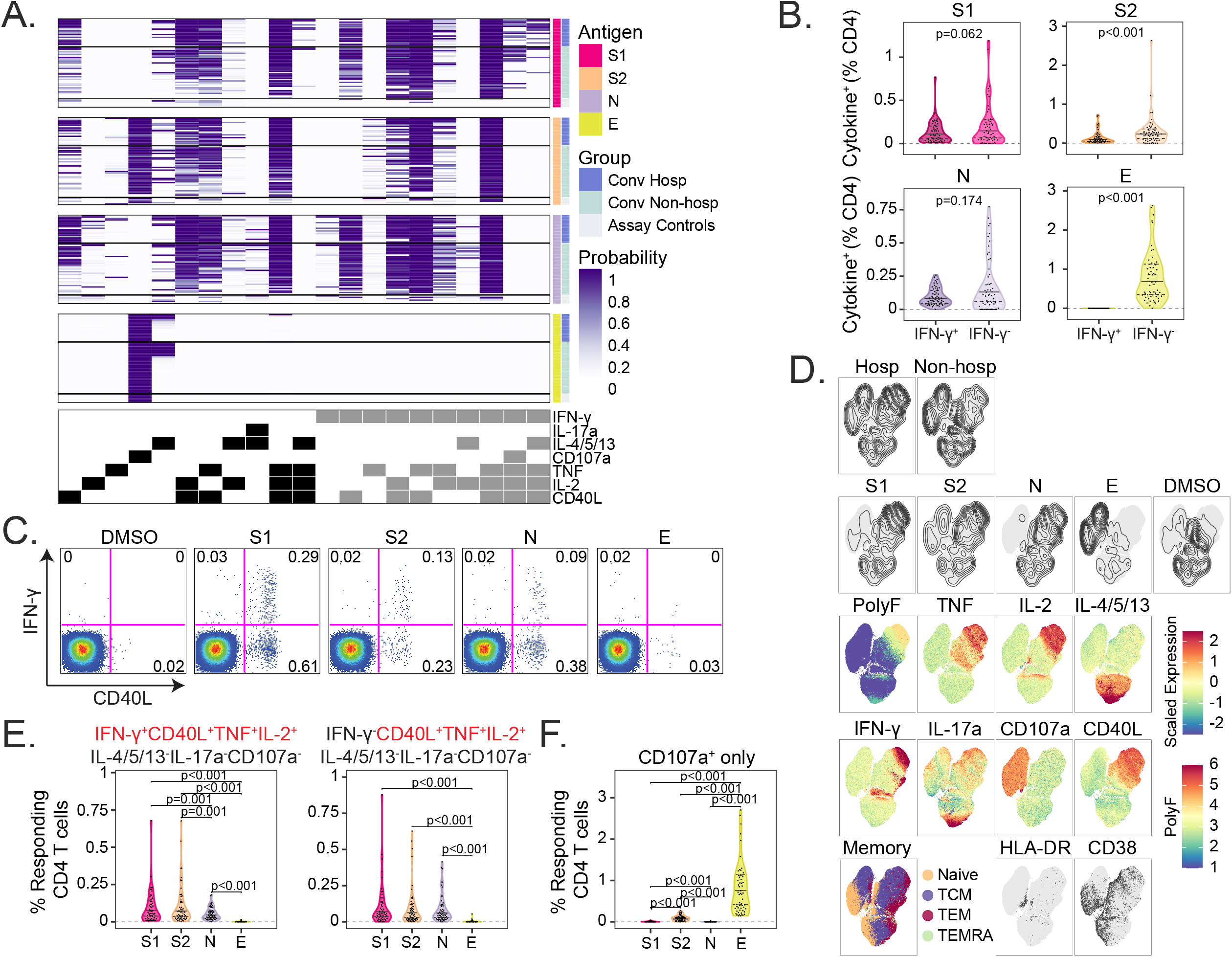
IFN-γ independent CD4 T-cell responses to SARS-CoV-2 structural antigens. (A) Intracellular cytokine staining (ICS) was used to profile the functions of CD4 T cells specific for the S1 and S2 domains of spike, nucleocapsid (N), and envelope small membrane protein (E). Data were analyzed using COMPASS, and results are displayed as a probability heatmap in which the rows represent study subjects and the columns represent CD4 T cell functional subsets. The depth of shading within the heatmap represents the probability of detecting a response above background. Responses are stratified by group to enable comparisons across stimulation conditions. In the column legend, white indicates absence and black/gray indicates presence of a function, respectively. (B) Background subtracted magnitudes of CD4+ T cell responses stratified by the presence of IFN-γ are compared between groups. (C) Representative bivariate flow cytometry plots showing the expression of IFN-γ and CD40L following stimulation with a negative control (DMSO), or peptide pools targeting S1, S2, N, and E. (D) Cells expressing any of the functional profiles identified by COMPASS were aggregated across all subjects prior to performing dimensionality reduction with uniform manifold approximation and projection (UMAP). Plots are stratified and colored according to the hospitalization status, stimulation (DMSO, S1, S2, N, and E), effector function (IFN-γ, TNF, IL-2, IL-4/5/13, IL17a, CD107a, and CD40L), memory markers (Naive: CD45RA+CCR7+; central memory (TCM): CD45RA-CCR7+; effector memory (TEM): CD45RA-CCR7-; and effector memory RA (TEMRA): CD45RA+CCR7-) and activation markers (HLA-DR, CD38). Mean fluorescence intensities (MFI) were scaled to achieve a mean of zero and standard deviation of one. Polyfunctionality (PolyF) was calculated as the number of cytokines gated positive for each cell. (E) Background corrected magnitudes of CD4+ T cells expressing a CD40L+IL-2+TNF+ functional profile in the presence or absence of IFN-γ are compared between groups for each stimulation. (F) Background corrected magnitudes of CD4+ T cells expressing CD107a in the absence of all other functions are compared between groups. Wilcoxon signed-rank tests were used to compare frequencies between groups in panels B and E. Correction for multiple hypothesis testing was achieved using the Bonferroni method for panel E, but panel B reports unadjusted p-values. Median, 25th, and 75th quartiles are indicated for violin plots. If not shown, p-values were not significantly different.

**Figure 4.**
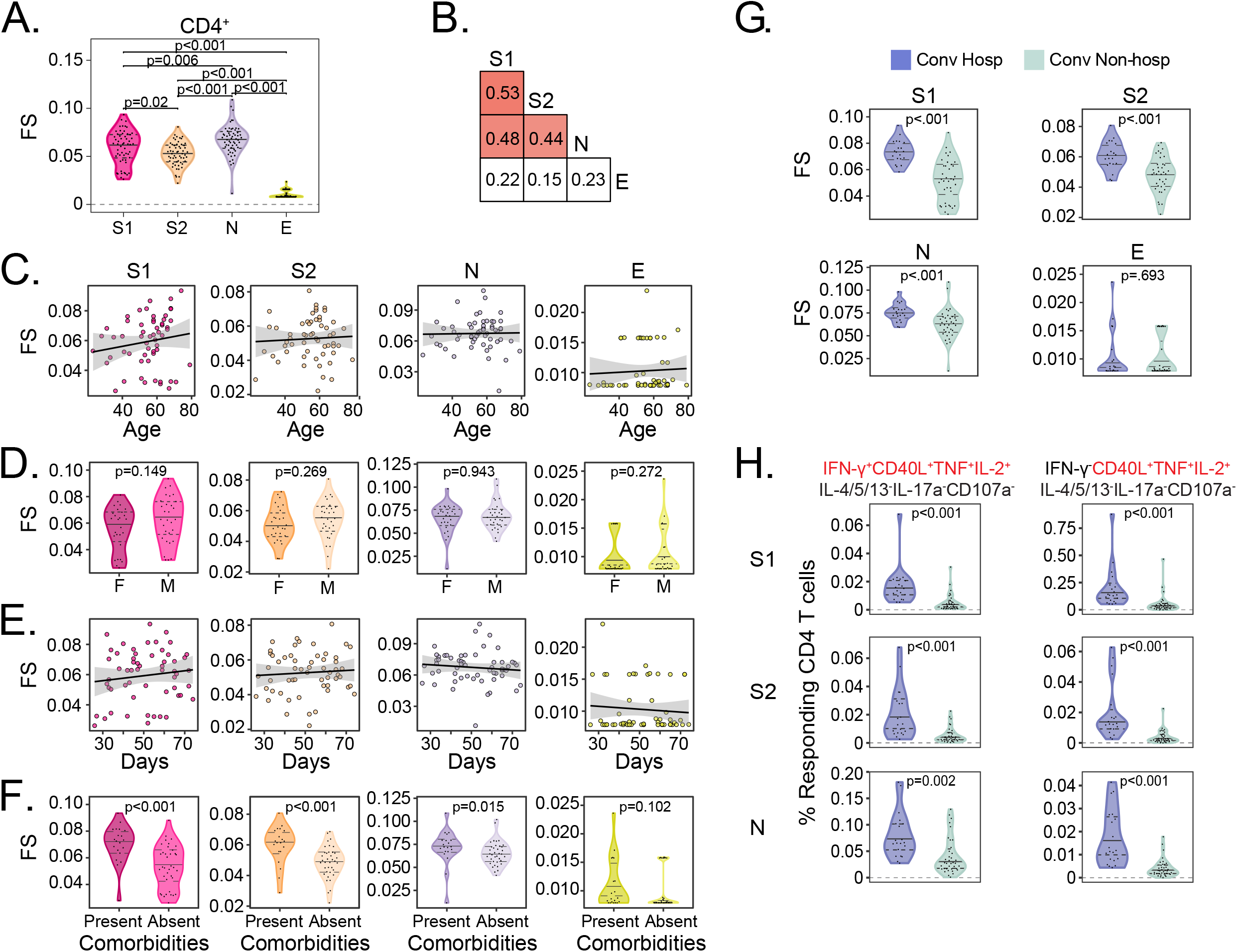
Functional diversity of CD4 T cell responses to SARS-CoV-2 are associated with hospitalization. (A) The CD4 T cell functionality score (FS) was determined by COMPASS and compared across all four stimulation conditions (S1, S2, N, and E). (B) Two-way correlations in functionality scores were compared between stimulation conditions. Colored squares indicated a statistically significant correlation (p < 0.05). For each stimulation, we examined the association with (C) Age, (D) Sex, and (E) days since symptom onset. The black lines on the scatter plots represent best fit linear regression lines, and the grey-shaded areas represent the 95% confidence interval of the predicted means. (F) CD4 functionality scores for each stimulation were compared in the presence and absence of comorbidities. (G) Background corrected magnitudes of CD4+ T cells expressing a CD40L+IL-2+TNF+ functional profile in the presence or absence of IFN-γ are compared between groups after stimulation with S1, S2, and N. CD4 functionality scores were compared using Wilcoxon signed-rank tests or Mann-Whitney U tests were used followed by correction for multiple hypothesis testing using the Bonferroni method except for panels D and F. Supplementary Figure 5 shows all the functional profiles that were compared to obtain p-values reported in panel G. Median, 25th, and 75th quartiles are indicated for violin plots. If not shown, p-values were not significantly different.

### IFN-γ independent CD4 T-cell responses to SARS-CoV-2 structural antigens

We next investigated the functional profiles of SARS-CoV-2 specific T cells. PBMCs were stimulated with overlapping peptide pools targeting the S1 or S2 domain of spike, nucleocapsid (N), or envelope small membrane protein (E). We used intracellular cytokine staining (ICS) to identify antigen-specific T cells expressing interleukin 2 (IL-2), IL-4/5/13, IL-17a, IFN-γ, tumor necrosis factor (TNF), CD107a, and CD40L (Supplementary Figure 3). To ensure the detection of polyfunctional T cell subsets that may be present at low frequencies, we employed COMbinatorial Polyfunctionality analysis of Antigen-Specific T cell Subsets (COMPASS)(40). Among 128 possible functional profiles, we detected 21 antigen-specific CD4 T cell subsets across all four peptide pool stimulations (Figure 3A). Notably, the probability of detecting a particular response varied according to the antigen. For example, several profiles containing three or four functions were readily detected after stimulation with S1, S2, or N but not E. However, the two profiles containing five functions (IFN-γ, IL-14/5/13, TNF, IL-2, and CD40L) were only detected after stimulation with S1. Stimulation with E resulted in a CD107a monofunctional profile that was also observed after stimulation with S2 (Figure 3A).

Notably, 11 (52%) of the 21 CD4 T cell functional profiles identified by COMPASS did not contain IFN-γ (Figure 3A). Because COMPASS only reports the probability of detecting a particular response, we next examined the magnitude of T cell responses stratified by the presence of IFN-γ. We found nearly equivalent numbers of IFN-γ^+^ and IFN-γ^-^ T cells after stimulation with S1 or N. However, more T cells expressed IFN-γ-independent functions after stimulation with S2 and E (Figure 3B and 3C). These data suggest that a substantial fraction of the SARS-CoV-2-specific T cell response could be missed by conventional assays, such as IFN-γ ELISPOT(41). We used uniform manifold approximation and projection (UMAP) to examine qualitative associations between hospitalization status, stimulation, and T cell functional profiles. Hospitalization appeared to be associated with responses to S1, S2, and N, though there was overlap with non-hospitalized subjects (Figure 3D). The degree of polyfunctionality appeared to be associated with hospitalization, which was also suggested by COMPASS (Figure 3A and 3D). Among the 21 functional profiles identified by COMPASS, CD4 T cells simultaneously expressing CD40L, IL-2, and TNF were detected at the greatest magnitudes, regardless of the presence of IFN-γ, and were highest after stimulation with S1 or S2 (Figure 3E and Supplementary Figure 5). By contrast, ∼1% of CD4 T cells expressed CD107a independent of IFN-γ after stimulation with E (Figure 3F). Finally, CD4 T cells with a detectable cytokine response predominantly expressed a CCR7+CD45RA-central memory phenotype, but very few demonstrated co-expression of the activation markers HLA-DR and CD38 (Figure 3D). Taken together, these data demonstrate the functional diversity of CD4 T cell responses to SARS-CoV-2 structural antigens driven in large part by IFN-γ-independent profiles that are not typically the focus of vaccine immunogenicity or epitope mapping studies (26, 42, 43).

**Figure 5.**
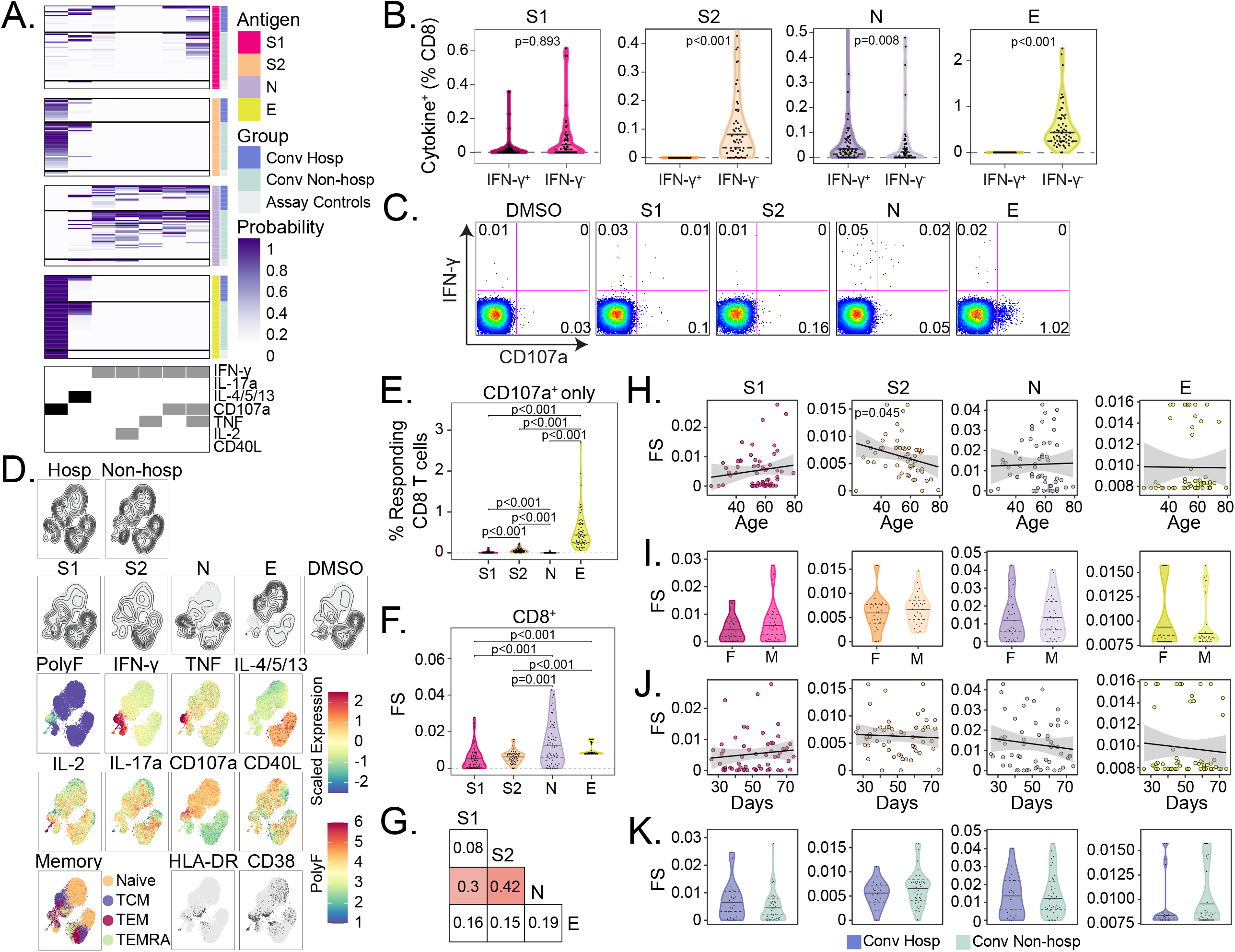
CD8 T cell responses to SARS-CoV-2 structural antigens are not associated with hospitalization. (A) Intracellular cytokine staining (ICS) was used to profile the functions of CD8 T cells specific for the S1 and S2 domains of spike, nucleocapsid (N), and envelope small membrane protein (E). Data were analyzed using COMPASS, and results are displayed as a probability heatmap in which the rows represent study subjects and the columns represent CD8 T cell functional subsets. The depth of shading within the heatmap represents the probability of detecting a response above background. Responses are stratified by group to enable comparisons across stimulation conditions. In the column legend, white indicates absence and black/gray indicates presence of a function, respectively. (B) Background subtracted magnitudes of CD8+ T cell responses stratified by the presence of IFN-γ are compared between groups. To facilitate visualization, a single outlier is not displayed for S2 and N. (C) Representative bivariate flow cytometry plots showing the expression of IFN-γ and CD107a following stimulation with a negative control (DMSO), or peptide pools targeting S1, S2, N, and E. (D) Cells expressing any of the functional profiles identified by COMPASS were aggregated across all subjects prior to performing dimensionality reduction with uniform manifold approximation and projection (UMAP). Plots are stratified and colored according to the hospitalization status, stimulation (DMSO, S1, S2, N, and E), effector function (IFN-γ, TNF, IL-2, IL-4/5/13, IL17a, CD107a, and CD40L), memory markers (Naive: CD45RA+CCR7+; central memory (TCM): CD45RA-CCR7+; effector memory (TEM): CD45RA-CCR7-; and effector memory RA (TEMRA): CD45RA+CCR7-) and activation markers (HLA-DR, CD38). Mean fluorescence intensities (MFI) were scaled to achieve a mean of zero and standard deviation of one. Polyfunctionality (PolyF) was calculated as the number of cytokines gated positive for each cell. (E) Background corrected magnitudes of CD8+ T cells expressing CD107a in the absence of all other functions are compared between groups. (F) The CD8 T cell functionality score (FS) was determined by COMPASS and compared across all four stimulation conditions (S1, S2, N, and E). (G) Two-way correlations in functionality scores were compared between stimulation conditions. Colored squares indicated a statistically significant correlation (p < 0.05). For each stimulation, we examined the association with (H) Age, (I) Sex, and (J) days since symptom onset. The black lines on the scatter plots represent best fit linear regression lines, and the grey-shaded areas represent the 95% confidence interval of the predicted means. Data were analyzed using Wilcoxon signed-rank tests (B, E, and F) and Mann-Whitney tests (I and K) and corrected for multiple hypothesis testing using the Bonferroni method except for panels I and K, which report unadjusted p-values. Median, 25th, and 75th quartiles are indicated for violin plots. If not shown, p-values were not significantly different.

### Functional diversity of CD4 T cell responses to SARS-CoV-2 are associated with hospitalization

Since UMAP revealed a qualitative association between T cell functional profile and hospitalization, we wanted to next explore that relationship quantitatively. To accomplish this, we used COMPASS to calculate a ‘functionality score’ (FS), which summarizes the functional breadth for each subject and stimulation into a continuous variable that can be incorporated into standard statistical models (40). Among CD4+ T cells, we found the highest functionality scores after stimulation with N, followed by S1, S2, then E (Figure 4A). However, the correlation between stimulations was modest, even between S1 and S2, confirming the importance of examining each antigen and functional domain independently (Figure 4B). CD4 functionality scores were not associated with age or sex for any of the antigens tested (Figure 4C and 4D). Notably, the functional breadth of CD4 T cell responses was stable over time (Figure 4E). Finally, we investigated whether functionality scores were associated with clinical risk factors and outcomes. We found higher functionality scores to S1, S2, and N but not E among hospitalized subjects and in the presence of medical comorbidities (Figure 4F and 4G). We examined this association using magnitudes of polyfunctional (CD40L+IL-2+TNF+) CD4 T cells and found the same to be true independent of the production of IFN-γ (Figure 4H). Thus, our data reveal that increased functional breadth of CD4+ T cell responses to spike and nucleocapsid are associated with known risk factors for severe COVID-19 independent of the production of IFN-γ.

### CD8 T cell responses to SARS-CoV-2 structural antigens are not associated with hospitalization

We next explored the functional breadth of CD8 T cell responses and its association with hospitalization. In contrast to the CD4 T cell response, COMPASS analysis identified seven T cell subsets, of which only two lacked IFN-γ (Figure 5A). IFN-γ independent T cell responses were dominant after stimulation with S2 and E (Figure 5A and 5B) and were characterized by expression of CD107a (Figure 5A and 5C). Both UMAP and COMPASS revealed polyfunctional profiles consisting of IFN-γ, IL-2, and TNF that were largely detected after stimulation with N in both hospitalized and not hospitalized subjects (Figure 5A and 5D). Similar to CD4 T cells, CD107a monofunctional CD8 T cells were mostly detected after stimulation with S2 and E (Figure 5E). Cytokine producing CD8 T cells were distributed across effector memory, central memory, and TEMRA phenotypes and did not co-express activation markers HLA-DR and CD38 (Figure 5D). Analysis of CD8 functionality scores revealed the greatest breadth after stimulation with N and very little correlation between antigens (Figure 5F and 5G). Again, we noted a surprisingly poor correlation between S1 and S2 that was driven by the dominance of polyfunctional responses to S1 and CD107a monofunctional responses to S2 (Figure 5A). Only S2 functionality scores were negatively correlated with age (Figure 5H). Finally, none of the stimulations were associated with sex, days post symptom onset, or hospitalization (Figure 5I, 5J, and 5K). Together, these data reveal that a thorough assessment of CD8 functional responses requires assays that examine more than IFN-γ, and that IFN-γ production and cytotoxic function are poorly correlated, even between the S1 and S2 domains of spike glycoprotein.

### Antigen-specific T cell and antibody responses are less coordinated among hospitalized subjects

Our results indicated a consistently higher magnitudes and increased functional breadth of several antibody and T cell features among hospitalized subjects. Thus, we next sought to identify the minimum set of features that could differentiate between hospitalized and non-hospitalized subjects. We used least absolute shrinkage and selection operator (LASSO) and identified eight features that consistently distinguished the two clinical groups via partial least squares discriminant analysis (PLS-DA) (Figure 6A and Supplementary Figure 6). With the exception of the induction of CD107a expression on NK cells by anti-RBD antibodies, all features were consistently enriched among hospitalized subjects (Figure 6B). When we examined the correlation between the selected features and all measured features, we noted that ADNP and FcR2A targeting spike were highly correlated with other features of humoral immunity (Figure 6C). Further, the four CD4 polyfunctional T cell features did not correlate with each other or with the humoral features, indicating a non-redundant contribution of T cell functions to the classification. Finally, we examined how T cell and antibody features correlated with each other in the two groups. Among non-hospitalized subjects, we noted more significant positive correlations between T cell and antibody features as compared to subjects who were hospitalized, even when the two groups were downsampled to account for the different sample sizes (Figure 6D and Supplementary Figure 7). These data suggest that non-hospitalized subjects are able to better coordinate antigen-specific T cells and antibody responses to SARS-CoV-2 despite having reduced functional breadth compared to subjects that were hospitalized.

**Figure 6.**
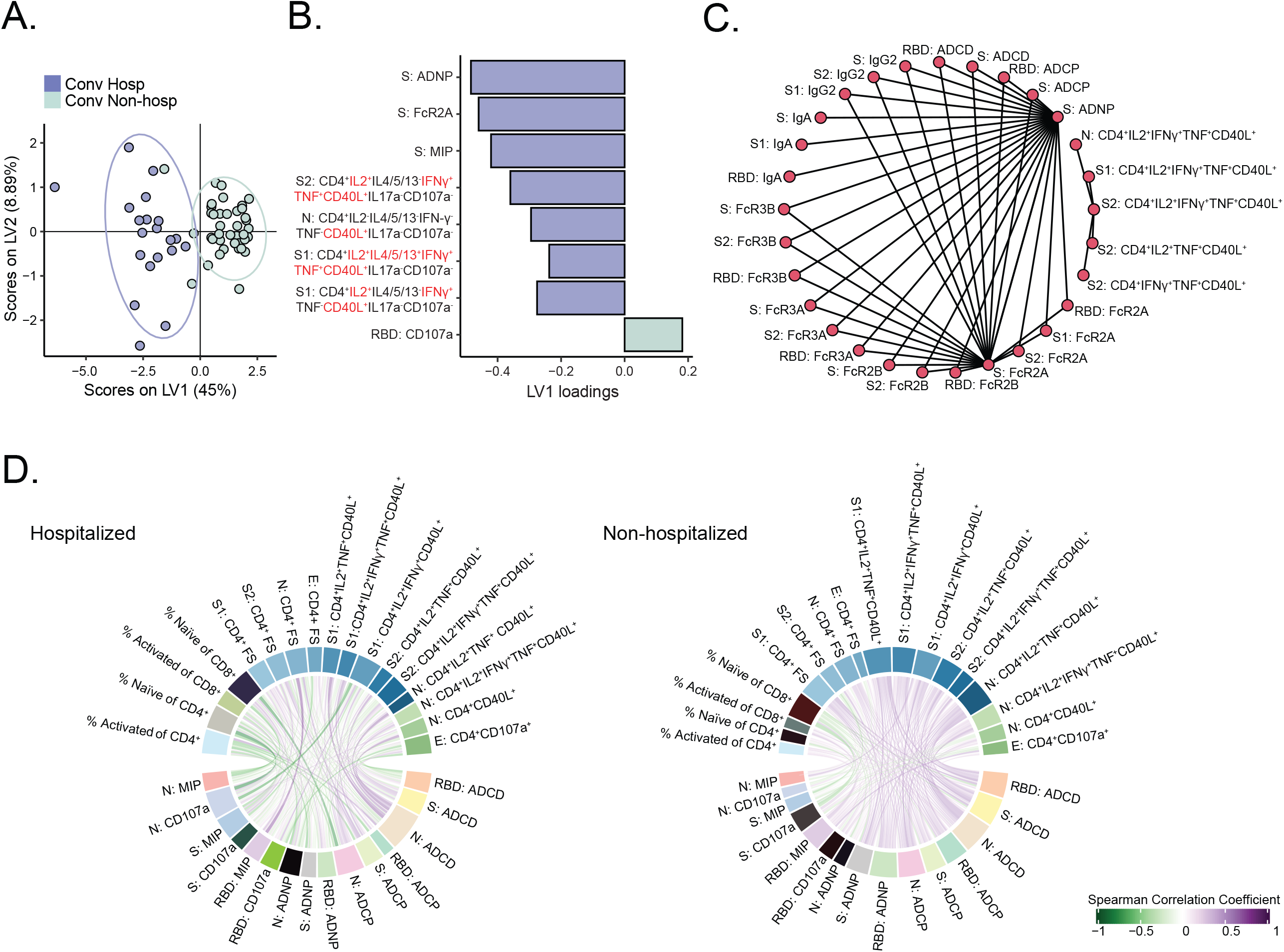
A classifier based on antibody and T cell features predicts hospitalization status. (A) Partial least squares discriminant analysis (PLS-DA) was used to identify T cell and antibody features that could discriminate between hospitalized (blue) and non-hospitalized (green) subjects. The PLS-DA scores plot shows the separation between the groups using the first two latent variables (LVs). Each dot represents an individual, and ellipses correspond to the 95% confidence regions for each group. (B) The bar plot shows the LV1 loadings of the LASSO-selected features for the PLS-DA ranked based on their Variable Importance in Projection (VIP) score. The features are color-coded according to the group in which they are enriched, i.e. the group with the higher average values of the feature. (C) The correlation network was generated from all the features correlated with LASSO-selected features. A cutoff with Spearman □ > 0.8 and p < 0.005 is shown. A cutoff of Spearman IZ > 0.8 with a Benjamini-Hochberg adjusted p-value < 0.05 was set and only connections outside of this cutoff are shown. The graph was generated using R package network(74, 76). (D) The chord diagram generated using the R package circlize(75) shows Spearman correlations between T cell features and antibody-dependent effector functions for non-hospitalized and hospitalized subjects, showing more positive correlations between these two immune system parts in the non-hospitalized group. Spearman correlations are shown as links that carry the color of the average correlation coefficient between the functional antibody features and T cell measurements. All correlations were visualized regardless of significance of correlation. The arc length of each segment is automatically scaled to the number of correlating segments it pairs with. To exclude potential bias caused by the number of subjects in non-hospitalized (n=40) and hospitalized (n=20) groups, per group Spearman correlations were calculated by sampling 10 subjects. This is repeated 100 times, and the average of the Spearman correlation coefficients were taken for each functional antibody feature - T cell measurement pair.

## DISCUSSION

In summary, we performed a cross-sectional study comprehensively examining the functional profiles of T cells and antibodies targeting SARS-CoV-2 spike, nucleocapsid, and envelope proteins in convalescent subjects who were either hospitalized or not hospitalized. We consistently found the magnitude and functional breadth of measured responses to be higher among hospitalized subjects and in the presence of medical comorbidities. However, these responses were more poorly correlated with each other when compared to non-hospitalized subjects. Since the presence of medical comorbidities are a known risk factor for severe disease and were over-represented among hospitalized subjects, these data support the possibility that virus-specific responses may contribute to immunopathology and severe COVID-19.

In contrast to most studies in which T cells or antibodies are studied in isolation, we comprehensively profiled both and analyzed them together in the context of detailed clinical information. In almost every respect, we find that they track together and show high levels of coordination among non-hospitalized subjects. The lack of coordination observed among hospitalized subjects may reflect a failure to control the virus at early stages, resulting in increased inflammation and virus load. Comorbid diseases were over-represented among hospitalized subjects, suggesting that they may be related to the increased functional breadth among T cells and antibodies that we describe here. Supporting this hypothesis are studies examining the effect of diabetes on adaptive immunity to *M. tuberculosis* (44). These studies have shown increased production of antigen-specific Th1 and Th17 cytokines in the presence of chronic hyperglycemia which is associated with an increased inflammatory state (45, 46). Whether SARS-CoV-2 specific T cell and antibody responses with increased functional breadth are the cause of poor clinical outcomes is not addressed by the cross-sectional design of our study and more definitively assessed in longitudinal studies or animal models.

Notably, we did not observe an association between neutralizing antibody titers and hospitalization in our study, which contrasts with emerging data examining patients much earlier in their disease course (47, 48). However, we did find that several functional attributes of spike-specific antibodies, including Ig subclass titers, were poorly correlated with neutralization yet associated with hospitalization. We have also previously shown that the ratio of spike:nucleocapsid antibodies is more predictive of death among hospitalized subjects than neutralization titers (37). These data add to a growing body of literature showing that several attributes of virus-specific antibodies are associated with clinical outcomes, including hospitalization (49). In general, we found that Ig subclass titers and Fc-specificity, and Fc-effector functions were lower among non-hospitalized subjects yet were more highly correlated with each other compared to hospitalized subjects. These findings may be the result of differences in innate immune activation, which may contribute to increased viral clearance and lower antigen loads. Innate immunity is known to be impaired in older subjects and in the presence of co-morbidities like diabetes (50).

We observed that T cell responses to envelope protein were qualitatively different from spike and nucleocapsid. Among both CD4 and CD8 T cells, a uniform functional profile of CD107a expression emerged, which was not seen with the other antigens. One potential explanation for this is that while E is abundantly expressed, very few molecules are incorporated into virions. Rather, E protein is mainly found in the endoplasmic reticulum-Golgi intermediate compartment (ERGIC) where it may readily access antigen-processing and presentation pathways (51, 52). Quantitatively, envelope dominated T cells responses, accounting for nearly ∼1% of all CD4 and CD8 T cells. In one study that examined genome-wide T cell responses to SARS-CoV-2 using activation-induced markers, envelope did not emerge as a prominent target (27). The reasons for this discordance are not clear but may be related to how antigen-specific T cells were identified. In our study, CD107a expression did not correlate with the activation-induced marker CD40L on CD4+ T cells, and several studies have shown that cytotoxic function correlates poorly with commonly examined surrogates, such as IFN-γ (53, 54).

Our data have several implications for the current race to develop a preventive vaccine for COVID-19. Phase I studies of subunit vaccines have quantified S-specific antibodies or neutralizing titers as well as IFN-γ production by S-specific T cells as evidence of immunogenicity (42, 43). However, we show that neutralizing antibody titers are poorly correlated with several important functional qualities of S-specific antibodies. We also show that a significant fraction of the CD4 T cell response to S does not include IFN-γ and depends on which domain is being examined. For example, CD4 and CD8 T cell responses to S2 were notable for having a cytotoxic phenotype compared to S1. In the integrated analysis, eight T cell and antibody features primarily focused on S1 were sufficient to classify hospitalized subjects with near perfect accuracy. These data raise the possibility that some vaccine-induced immune responses to spike glycoprotein might be harmful. Phase I studies that report safety are typically tested on young, healthy volunteers that are not representative of the target populations for candidate COVID vaccine, likely older and with medical comorbidities (55). This is a particularly important concern as several of the platforms being used, such as mRNA and adenoviral vectors, have limited experience in large clinical efficacy studies. An expanded analysis of the functions of vaccine-specific T cells and antibodies beyond what is required for regulatory approval will be required to understand the full benefits or risks of each approach.

## METHODS

### Study population

Whole blood samples were collected from individuals with laboratory-confirmed SARS-CoV-2 infection as part of a prospective longitudinal cohort study or as part of a protocol in support of expanded access to convalescent plasma for treatment of COVID-19 (ClinicalTrials.gov NCT04338360). Persons 18 years or older were eligible for inclusion 28 days or more after the resolution of symptoms. From the prospective study, individuals included in this report were from two groups: previously hospitalized inpatients and non-hospitalized outpatients. Inpatients were hospitalized at Harborview Medical Center, University of Washington Medical Center or at Northwest Hospital in Seattle, Washington and were identified through a laboratory alert system. Patients were enrolled during their hospital admission and had samples collected during their hospitalization. After hospital discharge, these participants were asked to present to an outpatient clinical research site approximately 30 days after symptom onset for follow-up. In person follow-up only occurred if participants were asymptomatic as per Center for Disease Control and Prevention (CDC) guidelines. Outpatients were identified through a laboratory alert system, email and flyer advertising, and through positive COVID-19 cases reported by the Seattle Flu Study (34). Outpatients completed their enrollment, data collection questionnaire, and first blood draw at an outpatient clinic visit approximately days 30 after symptom onset. All participants subsequently were asked to return at day 60 and then at day 90 or 120 for a subsequent follow-up. From protocol NCT04338360, only subjects with a history of hospitalization were considered for inclusion in this report. Sociodemographic and clinical data were collected from chart review and from participants at the time of enrollment (56), including information on the nature and duration of symptoms, medical comorbidities, and care-seeking behavior (Supplementary Table 1). Separately, assay control samples were derived from a 2017 adult specimen repository study or obtained from Bloodworks, Inc. (Seattle, WA).

### Study Approval

The studies were approved by the University of Washington Human Subjects Institutional Review Board, and all participants, or their legally authorized representatives, completed informed consent.

### Sample processing

All whole blood patient samples were collected in acid citrate dextrose or sodium heparin tubes (one subject) and immediately transferred to the University of Washington. Whole blood was centrifuged at 200xg for 10 minutes to separate plasma. Plasma was collected, centrifuged at 1200xg to remove debris, aliquoted, and stored at -20°C. Hank’s balanced salt solution (HBSS) (Thermo Fisher Scientific, Waltham, MA) or 1x phosphate buffered saline (PBS) (Thermo Fisher Scientific, Waltham, MA) was added to the whole blood cellular fraction to replace plasma volume. Peripheral blood mononuclear cells (PBMC) were isolated by density-gradient centrifugation using Histopaque (Sigma-Aldrich, St. Louis, MO). After washing, purified PBMC were resuspended in 90% heat-inactivated fetal bovine serum (FBS) (Sigma-Aldrich, St. Louis, MO) with 10% dimethyl sulfoxide (DMSO) (Sigma-Aldrich, St. Louis, MO) cryopreservation media and stored in liquid nitrogen until use. Both plasma and PBMC were frozen within six hours of collection time.

### Antibody neutralization

The SARS-CoV-2 pseudoviruses expressing a luciferase reporter gene were generated in an approach similar to as described previously (9, 10, 21). Briefly, the packaging plasmid psPAX2 (AIDS Resource and Reagent Program, Germantown, MD), luciferase reporter plasmid pLenti-CMV Puro-Luc (Addgene, Watertown, MA), and spike protein expressing pcDNA3.1-SARS CoV-2 SΔCT were co-transfected into HEK293T cells by lipofectamine 2,000 (Thermo Fisher Scientific, Waltham, MA). The supernatants containing the pseudotype viruses were collected 48 hours post-transfection, which were purified by centrifugation and filtration with a 0.45 µm filter. To determine the neutralization activity of the serum or plasma samples from cohorts, HEK293T-hACE2 cells were seeded in 96-well tissue culture plates at a density of 1.75 × 10^4^ cells/well overnight. Three-fold serial dilutions of heat inactivated (56°C for 30 minutes) serum or plasma samples were prepared and mixed with 50 µL of pseudovirus. The mixture was incubated at 37°C for 1 hour before adding to HEK293T-hACE2 cells. 48 hours after infection, cells were lysed in Steady-Glo Luciferase Assay (Promega, Madison, WI) according to the manufacturer’s instructions. SARS-CoV-2 neutralization titers were defined as the sample dilution at which a 50% reduction in relative light unit (RLU) was observed relative to the average of the virus control wells.

### Antibody titer measurements and FcR binding

In order to measure antigen-specific antibody subclass, isotype, and Fc-receptor (FcR) binding levels, a customized multiplexed Luminex assay was utilized, as previously described(57). This allows for relative quantification of antigen-specific humoral responses in a high-throughput manner and detection of different antigens at once. A panel of SARS-CoV-2 antigens including the full spike glycoprotein (S) (provided by Eric Fischer, Dana Farber), receptor binding domain (RBD) (Provided by Aaron Schmidt, Ragon Institute) nucleocapsid (N) (Aalto Bio Reagents, Dublin, Ireland), S1 (Sino Biological, Beijing, China) and S2 (Sino Biological, Beijing, China) were used. In brief, antigens were coupled to uniquely fluorescent magnetic carboxyl-modified microspheres (Luminex Corporation, Austin, TX) using 1-Ethyl-3- (3-dimethylaminopropyl) carbodiimide (EDC) (Thermo Fisher Scientific, Waltham, MA) and Sulfo-N-hydroxysuccinimide (NHS) (Thermo Fisher Scientific, Waltham, MA). Antigen-coupled microspheres were then blocked, washed, and incubated for 16 hours at 4°C while rocking at 700 rpm with diluted plasma samples (1:1,000 for Fc-receptors, 1:500 for IgG1, and 1:100 for all other readouts) to facilitate immune complex formation. The following day, plates were washed using an automated plate washer (Tecan, Männedorf, Zürich, Switzerland) with 0.1% BSA and 0.02% Tween-20. Antigen-specific antibody titers were detected with Phycoerythrin (PE)-coupled antibodies against IgG1, IgG2, IgG3, IgG4, IgA, and IgM (SouthernBiotech, Birmingham, AL). To measure antigen-specific Fc-receptor binding, biotinylated Fc-receptors (FcR2AH, 2B, 3AV, and 3B, Duke Protein Production facility) were coupled to PE and then added to immune-complexed beads to incubate for 1 hour at room temperature while shaking. Fluorescence was detected using an Intellicyt iQue with a PAA robot arm and analyzed using Forecyt software. The readout was mean fluorescence intensity (MFI) of PE. All experiments were performed in duplicate while operators were blinded to study group assignment, and all cases and controls were run at the same time to avoid batch effects.

### Functional antibody measurements

Bead-based assays were used to quantify antibody-dependent cellular phagocytosis (ADCP), antibody-dependent neutrophil phagocytosis (ADNP), and antibody-dependent complement deposition (ADCD), as previously described (58–60). Fluorescent neutravidin beads (red for ADCD, yellow for ADNP, and ADCP) (Thermo Fisher Scientific, Waltham, MA) were coupled to biotinylated SARS-CoV-2 antigens RBD, S, and N and incubated with diluted plasma (ADCP and ADNP 1:100, ADCD 1:10 dilution) for 2 hours at 37°C. For measuring monocyte phagocytosis, 2.5×10^4^ THP-1 cells (ATCC, Manassas, VA) were added per well and incubated for 16 hours at 37°C. For ADNP, Ammonium-Chloride-Potassium ACK lysis was performed on whole blood from healthy blood donors (MGH blood donor center), and 5×10^4^ cells were added per well and incubated for 1 hour at 37°C. Then, a PacBlue anti-CD66b detection antibody (clone G10F5) (RUO) (BioLegend, San Diego, CA) was used to stain neutrophils. To assess antibody-dependent complement deposition, lyophilized guinea pig complement (Cedarlane, Burlington, ON, Canada) was reconstituted and added to each well for 20 minutes at 37°C. Subsequently, a fluorescein (FITC)-conjugated goat IgG fraction to guinea pig complement C3 (MP Biomedicals, Santa Ana, CA) was added to detect C3 binding. Following fixation, sample acquisition was performed via flow cytometry (Intellicyt, iQue Screener plus) utilizing a robot arm (PAA), and analysis occurred using Forecyt software. A phagocytosis score was calculated for ADCP and ADNP as (percentage of bead-positive cells) × (MFI of bead-positive cells) divided by 10,000. ADCD was reported as MFI of FITC C3 deposition.

For the measurement of antibody-dependent natural killer (NK) cell activating functions, an ELISA-based surrogate-assay was employed as described previously(61). Briefly, plates were coated with 3 ug/mL of antigen (S, RBD and N), and samples were added at a 1:50 dilution and incubated for 2 hours at 37°C. NK cells were isolated the day prior via RosetteSep (STEM CELL Technologies, Vancouver, Canada) from healthy buffy coats (MGH blood donor center) and rested overnight in 1 ng/mL IL-15 (STEMCELL Technologies, Vancouver, Canada). 5×10^4^ NK cells were then added to the ELISA plates containing the immune complexes and incubated for 5 hours at 37°C in the presence of CD107a PE-Cy5 (clone H4A3) (BD Biosciences, San Jose, CA), GolgiStop (BD Biosciences, San Jose, CA), and BFA (Sigma-Aldrich, St. Louis, MO). Following the incubation, cells were fixed with Perm A (Life Technologies, Carlsbad, CA) and stained for surface markers with anti-CD16 APC-Cy7 (clone 3G8), anti-CD56 PE-Cy7 (clone B159), and anti-CD3 PacBlue (clone SP34-2) antibodies (BD Biosciences, San Jose, CA). Subsequently, cells were permeabilized using Perm B (Thermo Fisher Scientific, Waltham, MA), and intracellular cytokine staining with anti-IFN-γ FITC (clone 4S.B3) and anti-MIP-1β PE (clone D21-1351) (BD Biosciences, San Jose, CA) was performed. NK cells were defined as CD3-, CD16+ and CD56+. Data were reported as percentage of cells positive for CD107a, MIP-1β, or IFN-γ. All functional assays were performed in duplicate with two donors if applicable.

### Flow cytometry of T cells

PBMC samples were thawed in warm thaw media consisting of RPMI 1640 (Gibco, Waltham, MA) supplemented with 10% FBS (Hyclone, Logan, UT) (R10), and 2 uL/mL Benzonase (MilliporeSigma, Burlington, MA) sterile-filtered and centrifuged at 250xg for 10 minutes. The supernatant was decanted, and the viable cells were enumerated using the Guava easyCyte (MilliporeSigma, Burlington, MA) with guavaSoft 2.6 software. The cells were centrifuged at 250xg for 10 minutes and rested overnight at a density of 2 million cells/mL. The following day, the cells were enumerated using the Guava easyCyte and analyzed using two multiparameter flow cytometry assays.

For surface marker staining, PBMC were plated at a density of up to 4×10^6^ cells/well in a 96-well U-bottom plate and washed twice with PBS (Gibco, Waltham, MA). The cells were then stained with Fixable Green Live/Dead (Life Technologies, Carlsbad, CA) according to manufacturer’s instructions and incubated for 15 minutes at room temperature. Live/Dead staining and all following steps were performed in the dark. At the end of the incubation, cells were washed twice in PBS and blocked by incubating the cells at 4°C for 15 minutes in a 1:1 mixture of human serum (Valley Biomedical, Winchester, VA) and FACS buffer (PBS supplemented with 0.2% bovine serum albumin (BSA) (Sigma, St. Louis, MO) sterile-filtered). Cells were stained with anti-CCR7 (clone 150503) (BD Biosciences, San Jose, CA) in the presence of 50 nM dasatinib (Cayman Chemicals, Ann Arbor, MI) at 37°C for 30 minutes. At the end of the incubation, the cells were washed and resuspended in FACS buffer containing 50 nM dasatinib with MR1-5-(2-oxopropylideneamino)-6-D-ribitylaminouracil (5-OP-RU) and CD1d-□-Galactosylceramide (□-GalCer) tetramers (National Institutes of Health Tetramer Core Facility, Atlanta, GA) for 60 minutes at room temperature. Following the tetramer stain, the cells were washed twice in FACS buffer and stained at 4°C for 30 minutes with an antibody cocktail prepared in FACS buffer supplemented with 1 mM ascorbic acid and 0.05% sodium azide (62). Antibodies included anti-CD3 ECD (clone UCHT1) (Beckman Coulter, Brea, CA), anti-CD4 APC-H7 (clone L200), anti-CD8□ BB700 (clone RPA-T8), anti-CD38 BV605 (clone HB7), anti-CD45RA BUV737 (clone HI100), anti-HLA-DR BUV395 (clone G46-6) (BD Biosciences, San Jose, CA), anti-CD14 BV650 (clone M5E2), anti-CD19 BV785 (clone SJ25C1), anti-CD56 PE-Cy5 (clone HCD56), anti-TCR Vδ2 Alexa Fluor 700 (clone B6) (BioLegend, San Diego, CA), and anti-TCR γ/δ PE-Vio770 (clone 11F2) (Miltenyi Biotech, Auburn, CA). The samples were subsequently washed with FACS buffer and PBS. Cells were then fixed in 1% paraformaldehyde (Electron Microscopy Sciences, Hatfield, PA) and PBS solution for 15 minutes at 4°C, washed, and resuspended in PBS containing 2 mM ethylenediaminetetraacetic acid (EDTA) and stored at 4°C until acquisition. Samples were acquired on a BD LSRFortessa (BD Biosciences, San Jose, CA) equipped with a high-throughput sampler and configured with blue (488 nm), green (532 nm), red (628 nm), violet (405 nm), and ultraviolet (355 nm) lasers using standardized good clinical laboratory practice procedures to minimize variability of data generated.

For intracellular cytokine staining (ICS), we stimulated cells with overlapping peptide pools (15mers overlapping by 11 amino acids) targeting the S1 or S2 domains of spike glycoprotein, nucleocapsid, or envelope proteins (JPT Peptide Technologies, Acton, MA). The S1 pool spans the N-terminal amino acid residues (1-643 amino acids, 158 peptides) of spike glycoprotein, while the S2 pool spans the C-terminal amino acid residues (633-1273 amino acids, 157 peptides). Each peptide pool was reconstituted with 40 or 50 uL of pure DMSO (Sigma-Aldrich, St. Louis, MO) then diluted with PBS for a final concentration of 100 ug/mL in 16% DMSO/84% PBS or 20% DMSO/80% PBS. PBMC were plated at a density of up to 1×10^6^ cells/well in a 96-well U-bottom plate and stimulated with 1 ug/mL of each peptide in the pool or 0.25 ug/mL Staphylococcal Enterotoxin Type B (SEB) (List Biological Laboratories, Inc., Campbell, CA), or 0.2% DMSO (Sigma-Aldrich, St. Louis, MO). In addition to antigen, the stimulation cocktail consisted of 1 ug/mL anti-CD28/49d (BD Biosciences, San Jose, CA), 10 ug/mL Brefeldin A (BFA) (Sigma-Aldrich, St. Louis, MO), GolgiStop (BD Biosciences, San Jose, CA) prepared according to manufacturer’s instructions, anti-CD107a PE-Cy7 (clone H4A3) (BD Biosciences, San Jose, CA). The cells were stimulated for 6 hours at 37°C, after which EDTA (Sigma-Aldrich, St. Louis, MO) was added at a final concentration of 2mM. Samples were then stored at 4°C overnight. The following day, PBMC were washed twice with PBS then stained for 20 minutes at room temperature with Fixable Aqua viability dye (Life Technologies, Carlsbad, CA) prepared according to manufacturer’s instructions. A preparation of anti-CCR7 BV711 antibody (clone 150503) (BD Biosciences, San Jose, CA) in FACS buffer was centrifuged at 10,000xg for 5 minutes and then added to the cells for 30 minutes at 37°C. At the end of the incubation period, PBMC were washed twice with FACS buffer then incubated for 10 minutes at room temperature with 1x FACS Lyse (BD Biosciences, San Jose, CA). After lysis, the cells were washed with FACS buffer twice then permeabilized by incubating for 10 minutes at room temperature with 1x FACS Perm II (BD Biosciences, San Jose, CA). The PBMC were again washed twice with FACS buffer then stained with the remaining markers for 30 minutes at 4°C and then washed with FACS buffer: anti-CD3 ECD (clone UCHT1) (Beckman Coulter, Brea, CA), anti-CD4 APC-H7 (clone L200), anti-CD8β BB700 (clone 2ST8.5H7), anti-CD38 BV605 (clone HB7), anti-HLA-DR BUV395 (clone G46-6), anti-CD40L/CD154 PE-Cy5 (clone TRAP1), anti-CD45RA BUV737 (clone HI100), IFN-γ BV421 (clone B27), anti-TNF FITC (clone MAb11), anti-IL-2 PE (clone MQ1-17H12), anti-IL-4 APC (clone MP4-25D2) (BD Biosciences, San Jose, CA), anti-CD14 BV785 (clone M5E2), anti-CD19 BV785 (clone SJ25C1), anti-IL-5 APC (clone TRFK5), anti-IL-13 APC (clone JES10-5A2), and anti-IL-17a Alexa Fluor 700 (clone BL168) (BioLegend, San Diego, CA). Finally, samples were fixed with 1% paraformaldehyde (Electron Microscopy Solution, Hatfield, PA) and washed with PBS. They were then resuspended in PBS supplemented with EDTA at a final concentration of 2 mM and stored at 4°C until acquisition. For all flow cytometry experiments, study groups were evenly distributed in each batch and operators were not blinded to study group assignments.

### Statistics

#### Flow cytometry data analysis

Initial compensation, gating, and quality assessment of flow cytometry data was performed using FlowJo version 9.9.6 (FlowJo, TreeStar Inc, Ashland OR) for T cell data or Forecyt software (Intellicyt, Albuquerque, NM) for the antibody data. Representative gating trees for the surface marker panel and ICS data are shown in Supplementary Figure 3. The surface marker and ICS flow cytometry data were then processed using the OpenCyto framework in the R programming environment (63). Samples with poor viability defined on the basis of low CD3 counts (<10,000 cells) or low CD4 counts (<3,000 cells) were excluded from analysis. For the ICS panel, data from 20 convalescent hospitalized and 37 convalescent non-hospitalized subjects were ultimately analyzed. For the surface marker panel, data from 15 convalescent hospitalized and 36 convalescent non-hospitalized subjects were analyzed.

To achieve a comprehensive and unbiased analysis of the functional profiles of antigen-specific T cells, we used COMPASS(40). COMPASS uses a Bayesian hierarchical framework to model all observed cell subsets and select those most likely to have antigen-specific responses. Notably, COMPASS reports only the probability of detecting a particular T cell functional profile, rather than the absolute magnitude, which we calculated separately. For a given subject, COMPASS was also used to compute a functionality score that summarizes the entire functionality profile into a single continuous variable that can be used for standard statistical modeling (e.g. regression). For the data presented here, COMPASS was applied to each of the antigen stimulations separately for CD4+ and CD8+ T cells. Each one of the analyses was unbiased and considered all of the 128 possible boolean combinations of cytokine functions. Subjects with a high probability of response across many subsets were accordingly assigned a high functionality score. Magnitudes of T cell responses were calculated independent of COMPASS as the proportion of gated events in the stimulated condition minus the proportion of gated events in the unstimulated condition. Statistics were performed using background subtracted magnitudes, although data are plotted as the maximum of zero or this value. The R package ComplexHeatmap (64) was used to visualize COMPASS posterior probabilities of response. R packages corrplot and ggpubr, among others, were also use for analysis (65, 66).

Uniform Manifold Approximation and Projection (UMAP) was performed on all CD4+ or CD8+ events which were pre-selected from COMPASS-identified boolean subsets using the uwot package in R (67, 68), with the following parameters: spread = 9, min_dist = 0.02. The following markers were used in the UMAP analysis: CD3, CD4, CD8b, TNF, CD107a, CD154, IL-2, IL-17a, IL-4/5/13, IFN-γ, CD45RA, CCR7, CD38, and HLA-DR. Fluorescence intensities of each marker were scaled within each batch prior to UMAP.

All the raw flow cytometry data are available for download from [SOURCE] under study accessions. The code to complete flow cytometry data analyses, including COMPASS, can be found at https://github.com/seshadrilab/.

#### Integrated analysis of T cell and antibody functional profiles

Classification models were trained to discriminate subjects between hospitalized and non-hospitalized subjects using all the measured humoral and T-cell responses. Models were built with an approach similar to what we have previously published, using a combination of the least absolute shrinkage and selection operator (LASSO) for feature selection and then classification using partial least square discriminant analysis (PLS-DA) with the LASSO-selected features (37, 61). The set of model inputs comprised functional and biophysical humoral responses and T-cell responses to the SARS-CoV-2 antigens RBD, S, and N. Input data were scaled and centered. Missing values on T-cell responses were imputed using k-nearest neighbors. R package “DMwR” version 0.4.1 *knnImputation* function (69). Model robustness was assessed using five-fold cross-validation. For each cross-validation run, subjects were randomly stratified into five subsets ensuring that both groups were represented in each subset, with four subsets serving as the training set and the fifth as the test set. Each subset served as the test set once; therefore, each individual was in the test fold exactly once for each cross-validation run. For each test fold, LASSO-based feature selection was performed on logistic regression using the four subsets designated as the training set for that fold. Fold specific LASSO was repeated ten times and features, which are selected nine times out of ten, were identified as selected features. Using these selected features, a fold-specific PLS-DA was trained on training data for that fold. A set of predicted group labels were recorded for each subset. The first two latent variables (LVs) from a PLS-DA model trained on the LASSO-selected features were visualized. LVs are compound variables composed of the LASSO-selected features. For visualization, 95% confidence ellipses were calculated assuming a multivariate t distribution. Features were ordered according to their Variable Importance in Projection (VIP) score, a score which is higher for features that contribute more to the model. Analyses were performed using R version 4.0.2 (2020-06-22).

Significance of model performance was evaluated using “negative control” models of permuted data and randomly selected size-matched features. The repetitions of five-fold cross-validation generated a distribution of model classification accuracies. Corresponding model accuracy distributions were measured for two negative control models. The first approach consisted of permutation testing by randomly shuffling the group labels, within the cross-validation framework described above (i.e., a cross-validation framework matched to the actual model) (70). The second approach was to randomly select a set of features the same size as the LASSO-selected feature set. These control processes were repeated 100 times to generate a distribution of model accuracies observed in the context of permuted data and randomly selected, size-matched feature sets. The predicted group label for each subject was compared to the true group label to obtain a classification accuracy. Exact p-values were obtained as the tail probability of the true classification accuracy in the distribution of control model classification accuracies. Because one of the LASSO-selected features (ADNP Spike) was highly correlated with 54% of all features, we further assessed the performance of randomly selected features by selecting only from the remaining 46% features. Further, we additionally built an alternative model by excluding ADNP Spike to examine whether the separation between the groups would be achieved in the absence of this feature and to identify the strongest surrogate of ADNP Spike that can discriminate subjects between the two groups. These analyses were performed using R package “ropls” version 1.20.0 (71) and “glmnet” version 4.0.2 (72)..

Correlations were performed using Spearman method followed by Benjamini-Hochberg multiple correction (73). The co-correlate network was generated using R package “network” version 1.16.0 (74) and the chord diagram was generated using R package circlize version 0.4.10 (75).

## Supporting information

Equator Checklist

## Data Availability

All data referred to in the manuscript are available upon request.

## AUTHOR CONTRIBUTIONS

C.S., K.K.Q.Y., S.F., C.A., and G.A wrote the manuscript with contributions from all authors. H.Y.C., A.W., R.C., and D.M.K enrolled the clinical cohorts and facilitated access to blood and plasma samples. C.R.W. and J.K.L. facilitated sample selection and analyzed the demographic and clinical data. C.R.W, J.K.L, K.S. and N.F. facilitated subject enrollment, including collection and processing of the samples with assistance from E.D.L, M.S.A., K.K.Q.Y, and C.S. E.D.L., M.S.A, K.K.Q.Y, and C.S. designed and executed the T cell experiments and analyzed the data. S.F. and C.A. performed the antibody experiments and analyzed the data. M.T.S. analyzed T cell data and visualized T cell and antibody data. C.L., D.C., and D.L. facilitated computational analysis, including integrated analysis of T cell and antibody data.

## ACKNOWLEDGEMENTS

We thank Ariana Magedson, Dylan McDonald, Angela LeClair, and Miko Robertson for assistance with participant enrollment, and Pavitra Roychoudhury for extraction of viral load data. We thank Christopher L. McClurkan, Victoria L. Campbell, Lawrence Hemingway, and Maxwell Krist for specimen processing. We would also like to thank the Aaron Schmidt Lab including Blake Hauser, Tim Cardonna and Jared Feldman at the Ragon Institute for protein production efforts, as well as Eric Fischer from the Dana Farber Cancer institute. Finally, we would like to thank Bloodworks Northwest for whole blood assay controls for T cell studies.

We acknowledge support from the Ragon Institute of MGH, MIT, the Massachusetts Consortium on Pathogen Readiness (MassCPR to G.A.), the Bill & Melinda Gates Foundation (235730 to G.A. and INV-016575 to H.Y.C), NIAID (U19 AI35995 to G.A., R01-AI125189 to C.S., and Contract HHSN272201400049C to D.M.K.), the Doris Duke Charitable Foundation (20160103 to C.S.), and the U.S. Centers for Disease Control and Prevention (CK000490 to G.A.).

## SUPPLEMENTAL INFORMATION LEGENDS

**Supplementary Figure 1. Stability of SARS-CoV-2 specific antibody subclass levels over time**. Magnitudes of (A) spike (S) and (B) nucleocapsid (N) specific antibodies are plotted in hospitalized (purple) and non-hospitalized (green) subjects by days since symptom onset and stratified by immunoglobulin subclass (IgM, IgG1, IgG2, IgG3, IgG4, and IgA). Black lines on the scatter plots represent best fit linear regression lines, and the grey-shaded areas represent the 95% confidence interval of the predicted means. All p-values are not significant, indicating that the measured responses do not change over time.

**Supplementary Figure 2. Univariate analysis of antigen-specific antibody responses**. Violin plots show all features measured via the antigen-specific customized luminex assay. The readout is mean fluorescent intensity (MFI), indicating relative antibody titer. In each graph, MFI was compared between hospitalized (purple) and non-hospitalized (green) subjects using a Mann-Whitney test and unadjusted p-values are reported. In total, 50 variables were measured: (A) IgG1, IgG2, IgG3, IgG4, IgA, and IgM, and (B) FcR2A, FcR2B, FcR3A, and FcR3B against spike (S), receptor binding domain (RBD), and nucleocapsid (N) antigens.

**Supplementary Figure 3. Gating strategy for T cell flow cytometry**. (A) Data presented in Figure 1 were obtained using a 15-color multiparameter flow cytometry panel. Events were first isolated from a time gate, followed by singlets. Viable cells were identified, and then CD19 and CD14 markers were used to identify B cells and monocytes, respectively. Gating then proceeded from lymphocytes to a second singlet gate. From the second singlet gate, CD56 was used to identify natural killer cells. In parallel, CD3+ T cells from the singlet gate were further characterized using CD1d-L-Galactosylceramide (L-GalCer) and MR1-5-(2-oxopropyl phenylamino)-6-D-ribitylaminouracil (5-OP-RU) tetramers to identify invariant natural killer T cells and mucosal-associated invariant T cells, respectively, as well as activation markers (HLA-DR and CD38), and γδ T cells (Pan-γδ and Vδ2). In addition, CD3+ T cells were also examined for co-receptor usage with CD4 and CD8 markers. Finally, memory populations were separately gated for CD4+ and CD8+ cells using CD45RA and CCR7. (B) Data presented in Figures 3-5 were obtained using a 14-color multiparameter intracellular cytokine staining (ICS) flow cytometry panel. A time gate was applied to the events, and then viable CD3+ T cells were identified. CD14 and CD19 markers were used to exclude monocytes and B cells, and then a singlet gate was applied. Lymphocytes were then gated and analyzed for HLA-DR (activation), CD38 (activation), and CD4 and CD8 co-receptor expression. For CD4+ and CD8+ populations, cells were characterized for expression of IFN-γ (Th1), IL-2 (Th1), TNF (Th1), IL4/5/13 (Th2), IL-17 (Th17), CD40L (activation and B cell help), CD107a (degranulation), CD45RA (memory), and CCR7 (memory) expression.

**Supplementary Figure 4. Cell frequencies of donor-unrestricted T cells, B cells, monocytes, and natural killer cells**. Flow cytometric analysis of peripheral blood mononuclear cells (PBMC) was performed using a 15-color surface staining and phenotyping panel. (A) Frequencies and activation statuses of invariant natural killer T (iNKT) cells and mucosal-associated invariant T (MAIT) cells were compared between hospitalized (purple) and non-hospitalized (green) subjects. Frequencies are displayed as percent of total T cells, and activation is calculated as the percentage total iNKT or MAIT cells that co-expressed HLA-DR and CD38. (B) B cells (CD19+), monocytes (CD14+), and natural killer (NK) cell (CD3-CD56+) frequencies are shown as percent of live cells and are compared between groups. (C) The frequency of activated (HLADR+CD38+) γδ T cells is plotted against days since symptom onset for both hospitalized and non-hospitalized subjects. T cell frequencies were compared between groups using Mann-Whitney U tests, followed by correction for multiple hypothesis testing using the Bonferroni method. Median, 25th, and 75th quartiles are indicated in the violin plots. The black line on the scatter plot represents a best fit linear regression line, and the grey-shaded area represents the 95% confidence interval of the predicted mean. If not shown, p-values were not significantly different.

**Supplementary Figure 5. Convalescent COVID-19 subjects demonstrate both IFN-**γ **dependent and independent CD4+ T cell responses following stimulation with SARS-CoV-2 protein antigens**. Background subtracted magnitudes of responding CD4 T cells is displayed for each of the functional subsets identified by COMPASS in Figure 3A after stimulation with peptide pools targeting (A) S1, (B) S2, (C) nucleocapsid, and (D) envelope. Boxplots indicating median and interquartile range are shown for hospitalized (purple) and non-hospitalized (green) subjects. Cell frequencies were compared between groups using the Mann-Whitney U tests followed by correction for multiple hypothesis testing using the Bonferroni method. Only significant p-values are indicated.

**Supplementary Figure 6. Validation of PLS-DA Model**. The classification accuracy distributions of the model presented in Figure 6 was compared to negative control models based on randomly selected or permuted data, by measuring the classification accuracies of each model in a five-fold cross-validation framework. (A) The violin plot shows the distributions of these classification accuracies for all three models across cross-validation replicates. Model performs significantly better compared to permuted labels. The model is not able to outperform the randomly selected features because a substantial portion of the measured features (54%) are significantly correlated (Spearman correlations, BH adjusted p-value < 0.05) with a LASSO-selected feature, ADNP Spike, thus are replaceable with ADNP Spike. (B) Features that are correlated with ADNP Spike were excluded. The model performs significantly better compared to randomly selected features from the pool of features, which are not significantly correlated with ADNP spike.

**Supplementary Figure 7. Correlations between the antibody and T-cell features are robust to sample size**. Heatmaps show Spearman correlations of antibody functions (in rows) with T-cell responses (in columns) using 40 non-hospitalized and 20 hospitalized subjects (A); the color of each cell is associated with the correlation coefficient and the significance of correlation is denoted with stars (*p<0.5, **p<0.01, ***p<0.001). To exclude potential bias caused by the number of subjects, 10 subjects were sampled per group. This was repeated 100 times, and the average of the Spearman correlation coefficient were taken for each functional antibody feature - T cell measurement pair. The color of each cell is associated with the average correlation coefficient and the numbers in the cell denote the number of times the correlation was significant (p < 0.05).

**Supplementary Table 1. Clinical and demographic features of the Clinical Cohort**. Raw data used to generate the values shown in Table 1 are included here. Demographics, clinical features, and viral loads (Ct) where available are shown for each individual.

