## Supplemental Figure 1 for "T cell and antibody functional correlates of severe COVID-19"

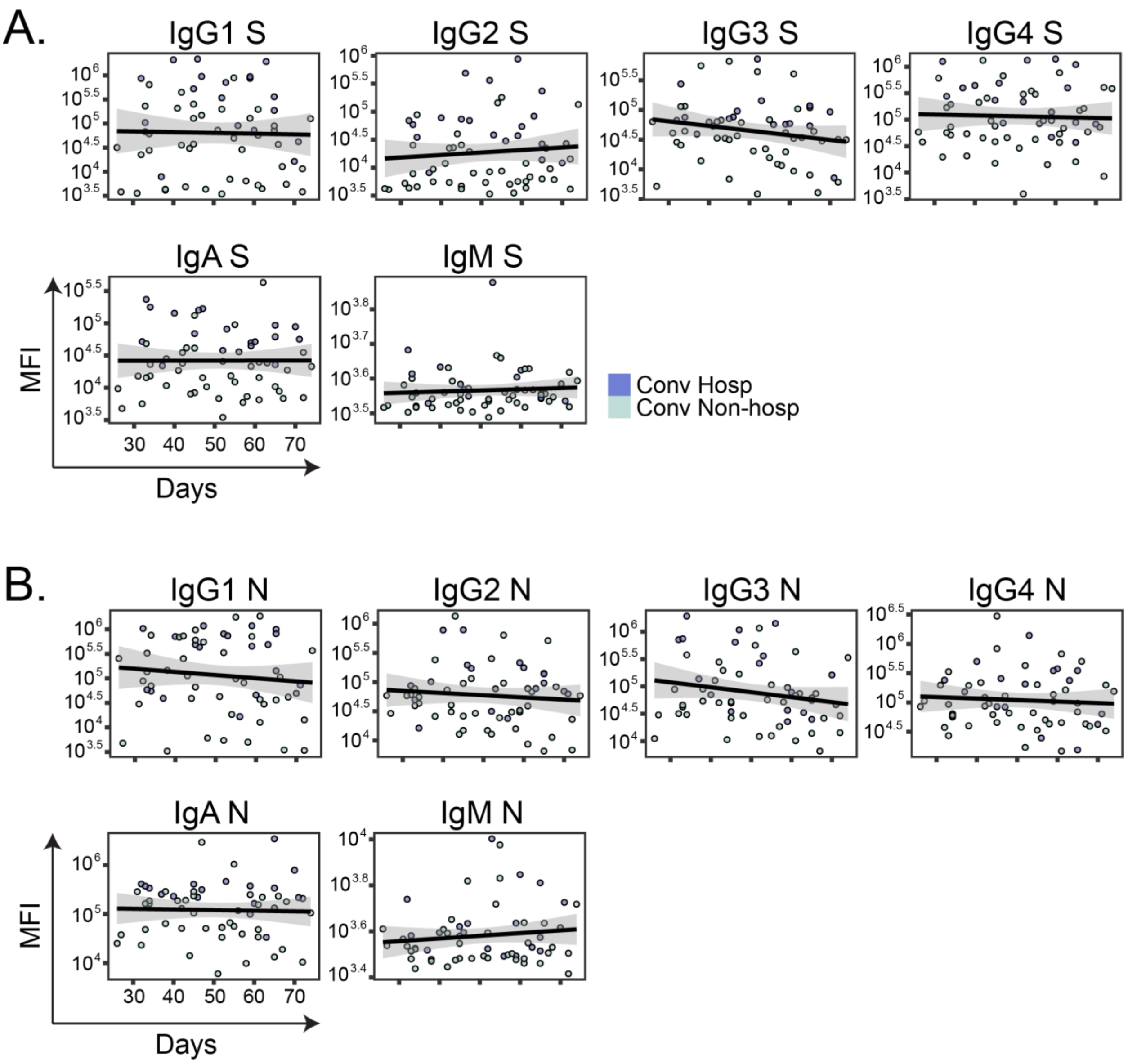


**Supplementary Figure 1. Stability of SARS-CoV-2 specific antibody subclass levels over time.** Magnitudes of (A) spike (S) and (B) nucleocapsid (N) specific antibodies are plotted in hospitalized (purple) and non-hospitalized (green) subjects by days since symptom onset and stratified by immunoglobulin subclass (IgM, IgG1, IgG2, IgG3, IgG4, and IgA). Black lines on the scatter plots represent best fit linear regression lines, and the grey-shaded areas represent the 95% confidence interval of the predicted means. All p-values are not significant, indicating that the measured responses do not change over time.
