## Supplemental Figure 2 for "T cell and antibody functional correlates of severe COVID-19"

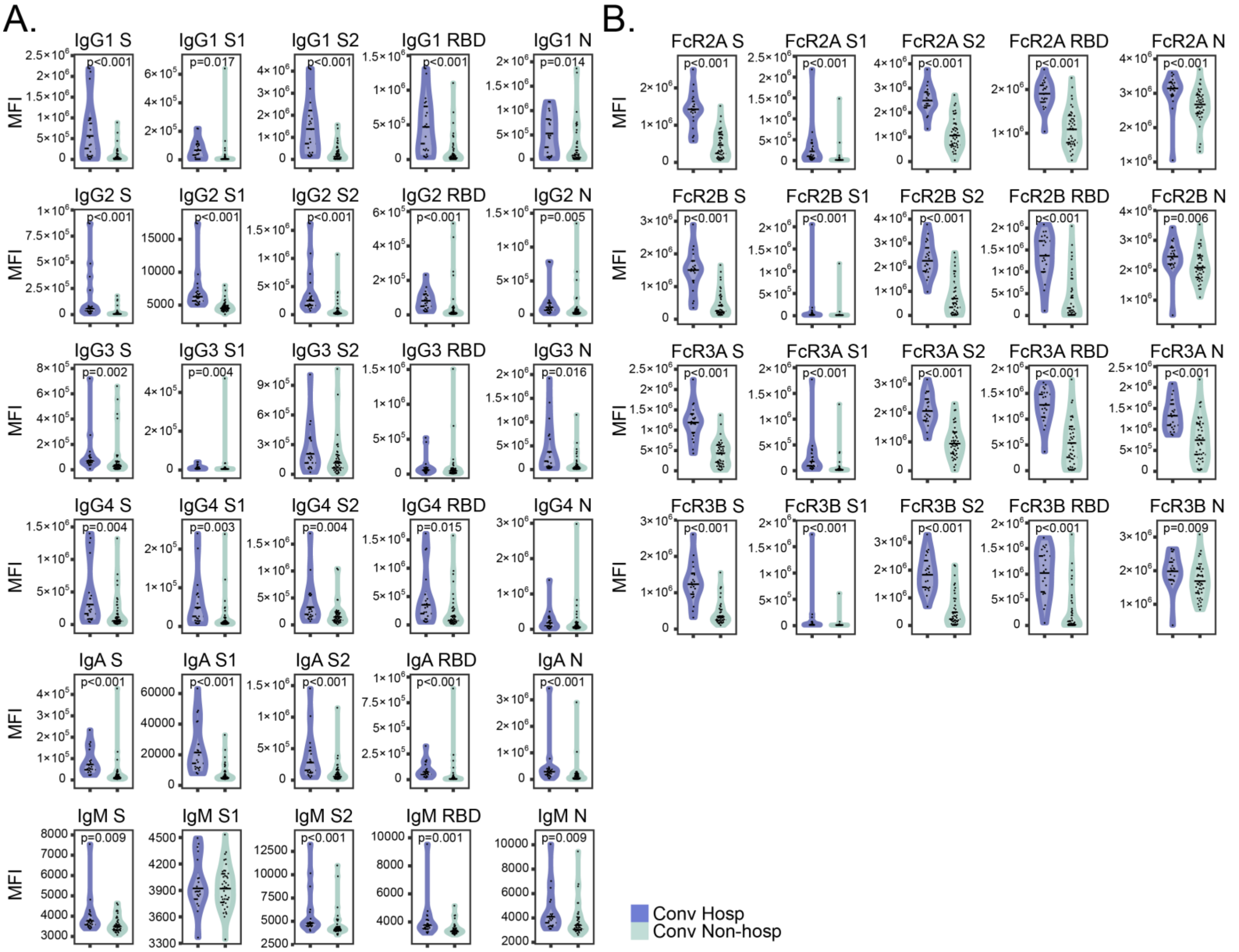


**Supplementary Figure 2. Univariate analysis of antigen-specific antibody responses.** Violin plots show all features measured via the antigen-specific customized Luminex assay. The readout is mean fluorescent intensity (MFI), indicating relative antibody titer. In each graph, MFI was compared between hospitalized (purple) and non-hospitalized (green) subjects using a Mann-Whitney test and unadjusted p-values are reported. In total, 50 variables were measured: (A) IgG1, IgG2, IgG3, IgG4, IgA, and IgM, and (B) FcR2A, FcR2B, FcR3A, and FcR3B against spike (S), receptor binding domain (RBD), and nucleocapsid (N) antigens.
