## Supplemental Figure 3 for "T cell and antibody functional correlates of severe COVID-19"

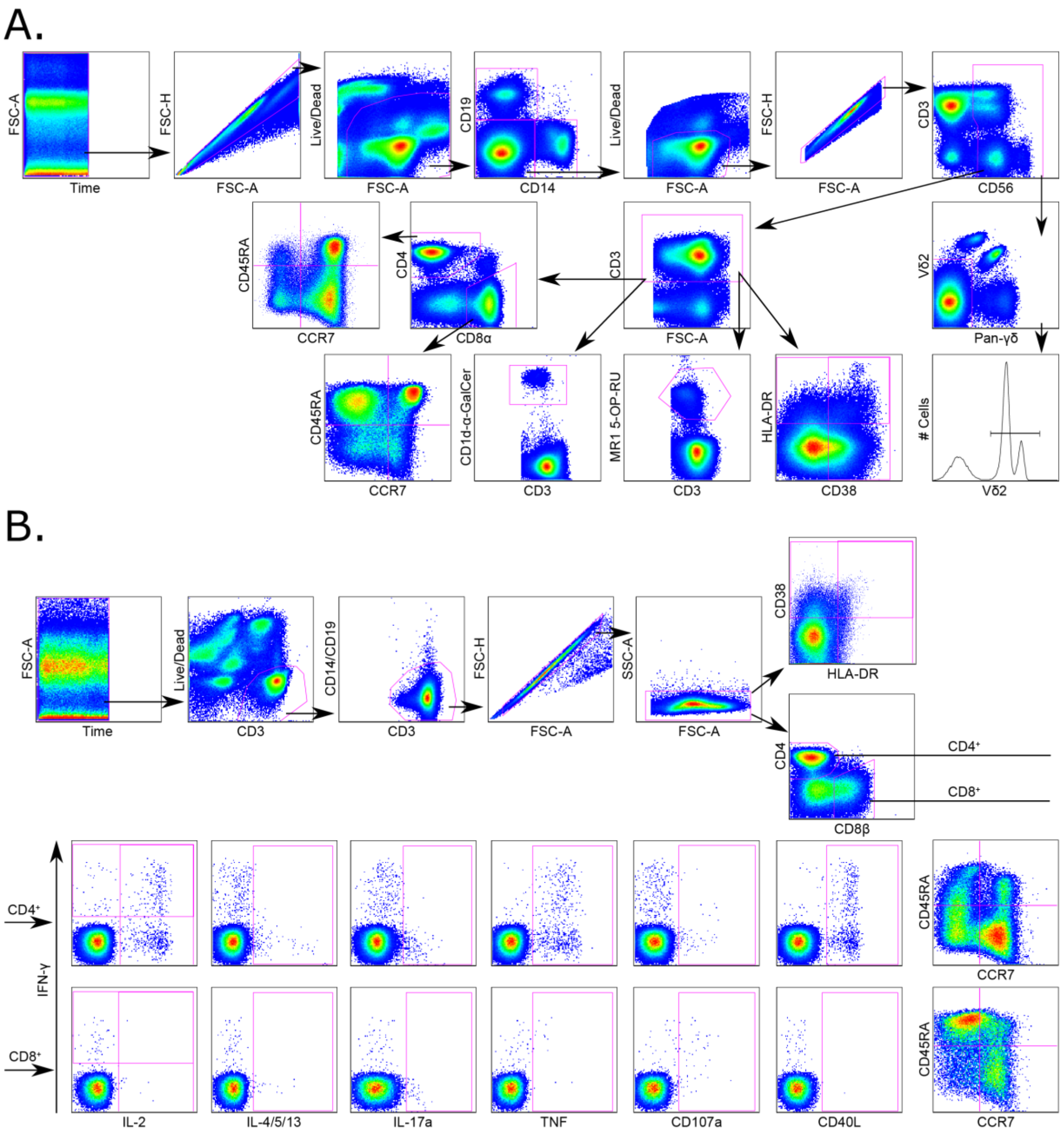


**Supplementary Figure 3. Gating strategy for T cell flow cytometry.** (A) Data presented in Figure 1 were obtained using a 15-color multiparameter flow cytometry panel. Events were first isolated from a time gate, followed by singlets. Viable cells were identified, and then CD19 and CD14 markers were used to identify B cells and monocytes, respectively. Gating then proceeded from lymphocytes to a second singlet gate. From the second singlet gate, CD56 was used to identify natural killer cells. In parallel, CD3+ T cells from the singlet gate were further characterized using CD1d-α-Galactosylceramide (α-GalCer) and MR1-5-(2-oxopropyl phenylamino)-6-D-ribitylaminouracil (5-OP-RU) tetramers to identify invariant natural killer T cells and mucosal-associated invariant T cells, respectively, as well as activation markers (HLA-DR and CD38), and γδ T cells (Pan-γδ and Vδ2). In addition, CD3+ T cells were also examined for co-receptor usage with CD4 and CD8 markers. Finally, memory populations were separately gated for CD4+ and CD8+ cells using CD45RA and CCR7. (B) Data presented in Figures 3-5 were obtained using a 14-color multiparameter intracellular cytokine staining (ICS) flow cytometry panel. A time gate was applied to the events, and then viable CD3+ T cells were identified. CD14 and CD19 markers were used to exclude monocytes and B cells, and then a singlet gate was applied. Lymphocytes were then gated and analyzed for HLA-DR (activation), CD38 (activation), and CD4 and CD8 co-receptor expression. For CD4+ and CD8+ populations, cells were characterized for expression of IFN-γ (Th1), IL-2 (Th1), TNF (Th1), IL4/5/13 (Th2), IL-17 (Th17), CD40L (activation and B cell help), CD107a (degranulation), CD45RA (memory), and CCR7 (memory) expression.
