## Supplemental Figure 4 for "T cell and antibody functional correlates of severe COVID-19"

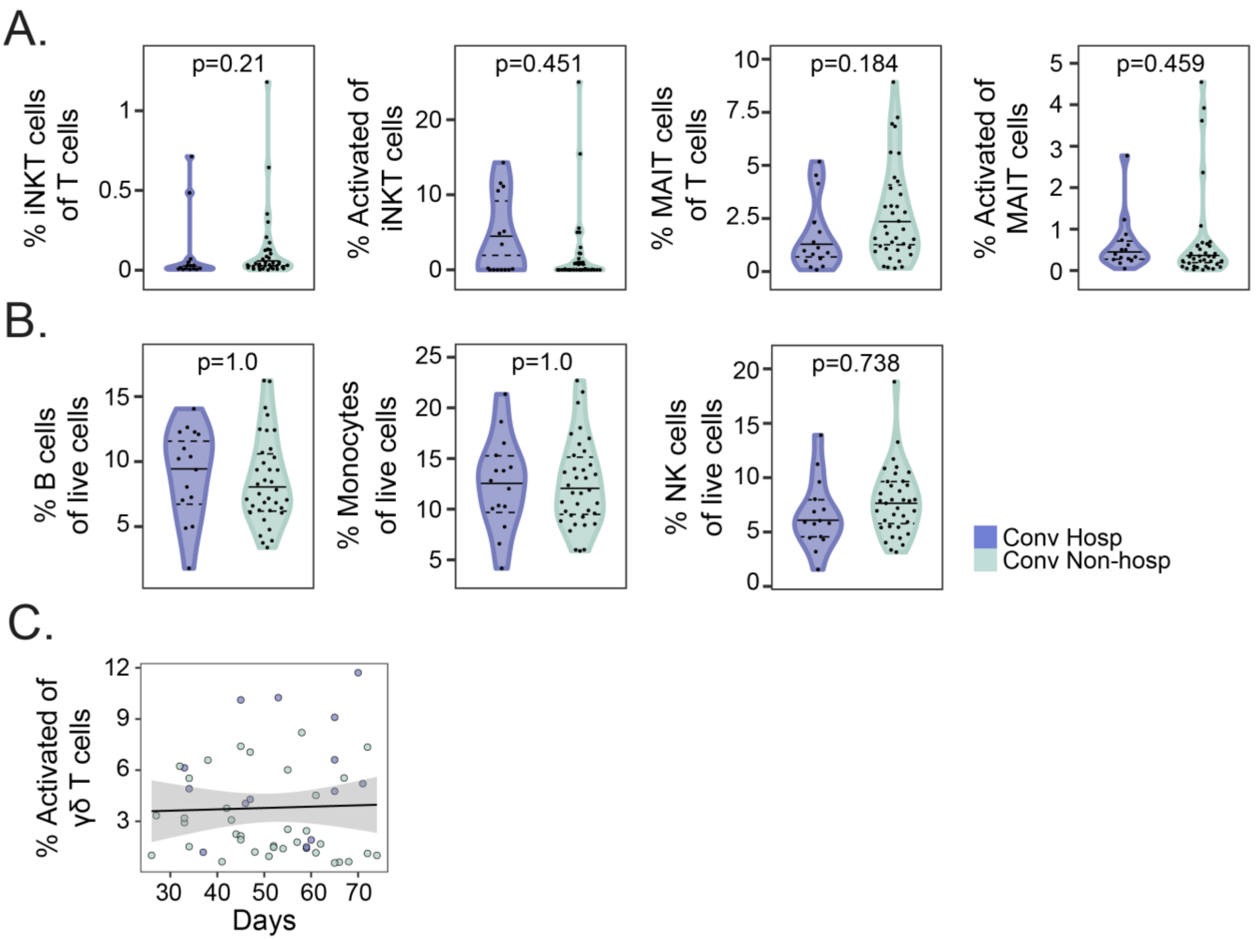


**Supplementary Figure 4. Cell frequencies of donor-unrestricted T cells, B cells, monocytes, and natural killer cells.** Flow cytometric analysis of peripheral blood mononuclear cells (PBMC) was performed using a 15-color surface staining and phenotyping panel. (A) Frequencies and activation statuses of invariant natural killer T (iNKT) cells and mucosal-associated invariant T (MAIT) cells were compared between hospitalized (purple) and non-hospitalized (green) subjects. Frequencies are displayed as percent of total T cells, and activation is calculated as the percentage total iNKT or MAIT cells that co-expressed HLA-DR and CD38. (B) B cells (CD19+), monocytes (CD14+), and natural killer (NK) cell (CD3-CD56+) frequencies are shown as percent of live cells and are compared between groups. (C) The frequency of activated (HLADR+CD38+) γδ T cells is plotted against days since symptom onset for both hospitalized and non-hospitalized subjects. T cell frequencies were compared between groups using Mann-Whitney U tests, followed by correction for multiple hypothesis testing using the Bonferroni method. Median, 25th, and 75th quartiles are indicated in the violin plots. The black line on the scatter plot represents a best fit linear regression line, and the grey-shaded area represents the 95% confidence interval of the predicted mean. If not shown, p-values were not significantly different.
