## Supplemental Figure 5 for "T cell and antibody functional correlates of severe COVID-19"

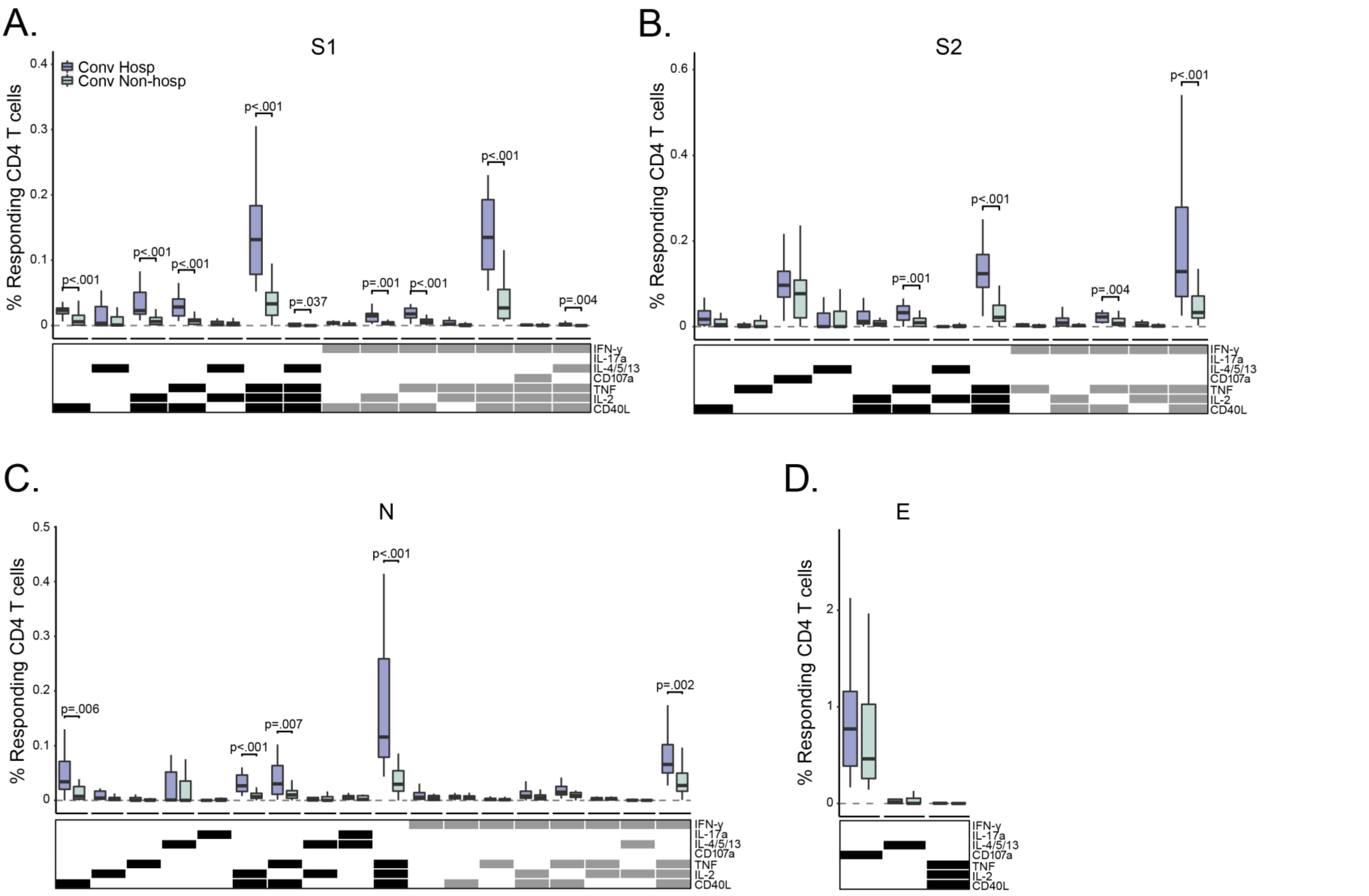


**Supplementary Figure 5. Convalescent COVID-19 subjects demonstrate both IFN-γ dependent and independent CD4+ T cell responses following stimulation with SARS-CoV-2 protein antigens.** Background subtracted magnitudes of responding CD4 T cells is displayed for each of the functional subsets identified by COMPASS in Figure 3A after stimulation with peptide pools targeting (A) S1, (B) S2, (C) nucleocapsid, and (D) envelope. Boxplots indicating median and interquartile range are shown for hospitalized (purple) and non-hospitalized (green) subjects. Cell frequencies were compared between groups using the Mann-Whitney U tests followed by correction for multiple hypothesis testing using the Bonferroni method. Only significant p-values are indicated.
