## Supplemental Figure 6 for "T cell and antibody functional correlates of severe COVID-19"

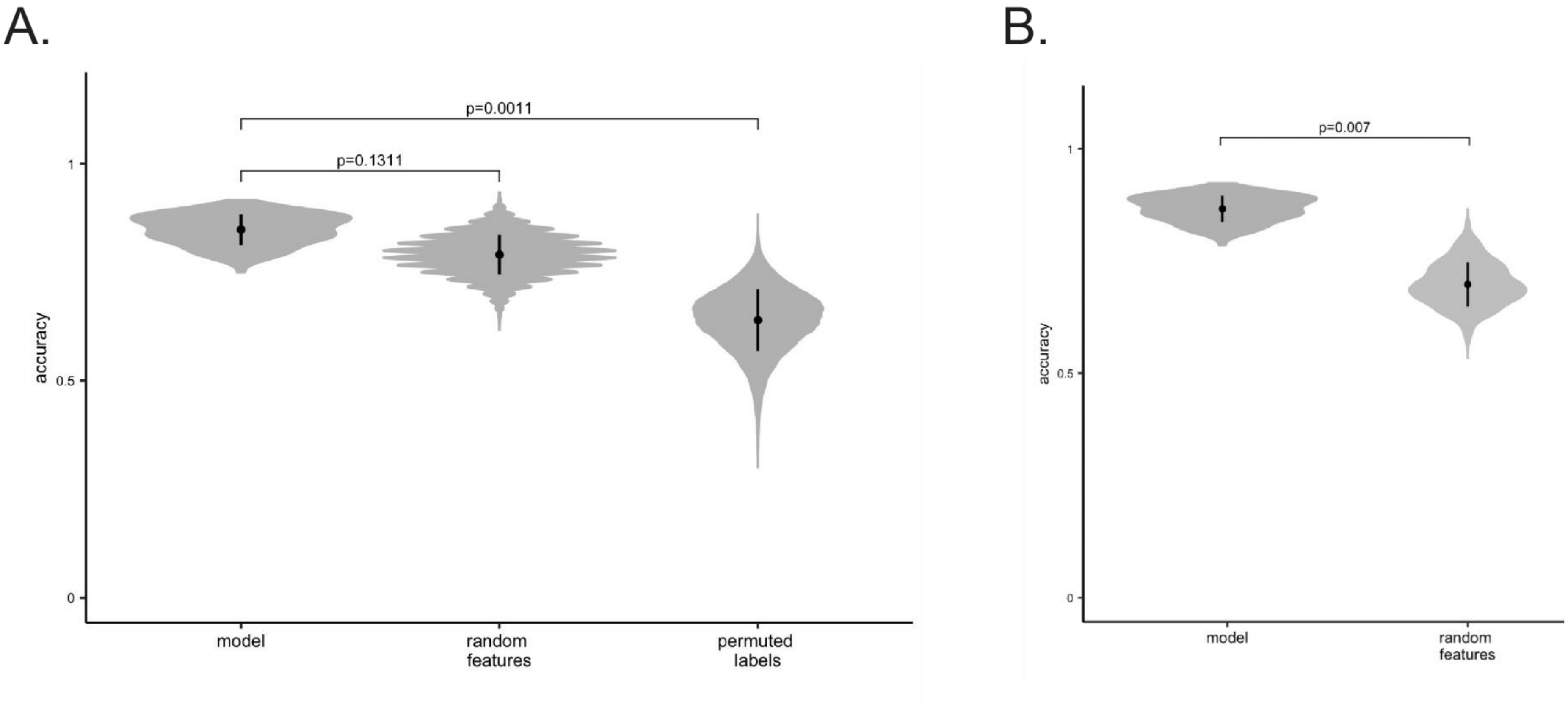


**Supplementary Figure 6. Validation of PLS-DA Model.** The classification accuracy distributions of the model presented in Figure 6 was compared to negative control models based on randomly selected or permuted data, by measuring the classification accuracies of each model in a five-fold cross-validation framework. (A) The violin plot shows the distributions of these classification accuracies for all three models across cross-validation replicates. Model performs significantly better compared to permuted labels. The model is not able to outperform the randomly selected features because a substantial portion of the measured features (54%) are significantly correlated (Spearman correlations, BH adjusted p-value < 0.05) with a LASSO-selected feature, ADNP Spike, thus are replaceable with ADNP Spike. (B) Features that are correlated with ADNP Spike were excluded. The model performs significantly better compared to randomly selected features from the pool of features, which are not significantly correlated with ADNP spike.
