## Supplemental Figure 7 for "T cell and antibody functional correlates of severe COVID-19"

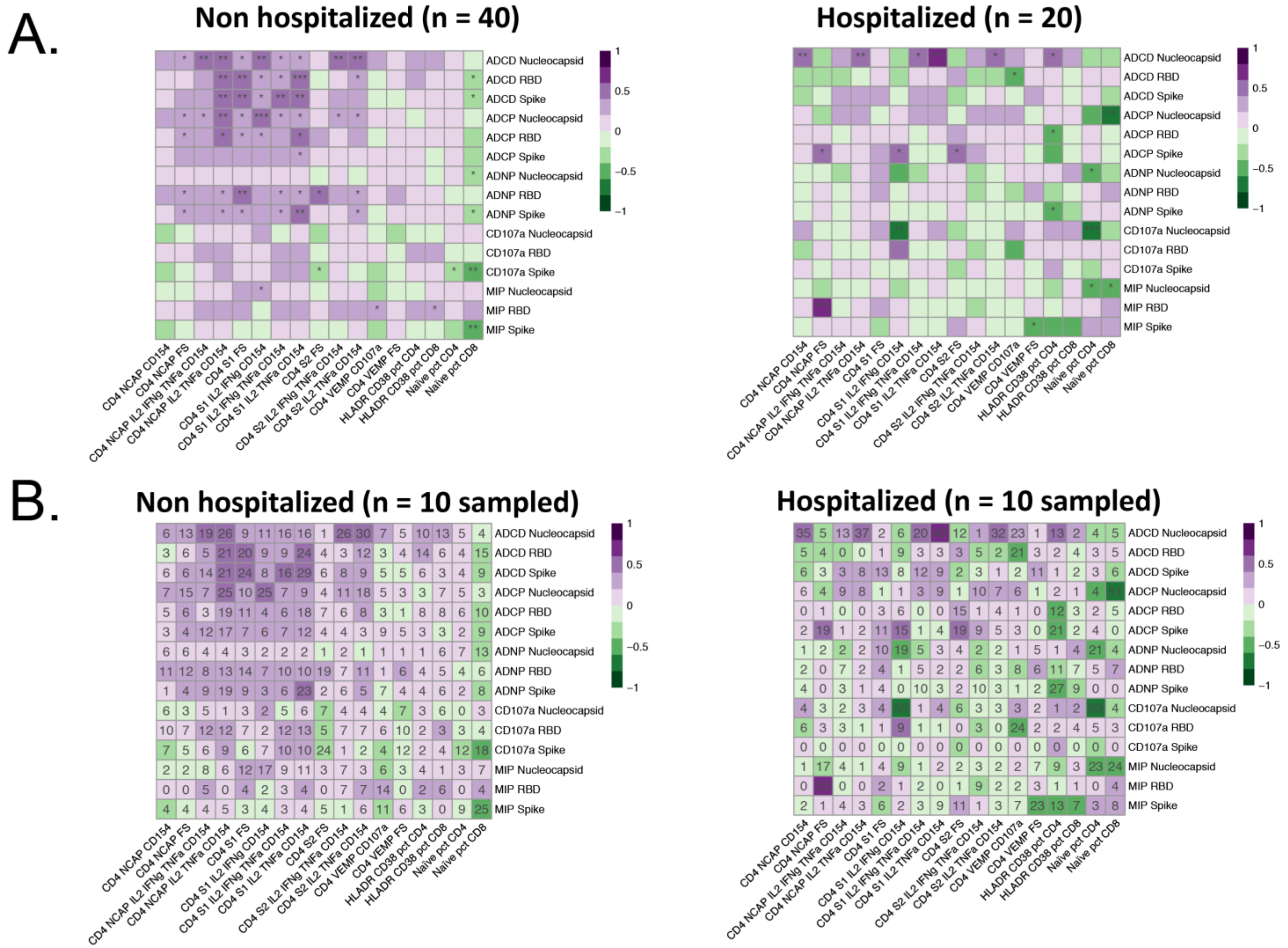


**Supplementary Figure 7. Correlations between the antibody and T-cell features are robust to sample size.** Heatmaps show Spearman correlations of antibody functions (in rows) with T-cell responses (in columns) using 40 non-hospitalized and 20 hospitalized subjects (A); the color of each cell is associated with the correlation coefficient and the significance of correlation is denoted with stars (*p<0.5, **p<0.01, ***p<0.001). To exclude potential bias caused by the number of subjects, 10 subjects were sampled per group. This was repeated 100 times, and the average of the Spearman correlation coefficient were taken for each functional antibody feature - T cell measurement pair. The color of each cell is associated with the average correlation coefficient and the numbers in the cell denote the number of times the correlation was significant ($p < 0.05$).
